# Harness Behavioural Analysis for Unpacking the Bio-Interpretability of Pathology Foundation Models

**DOI:** 10.64898/2025.12.31.25343151

**Authors:** Yang Hu, George Batchkala, Kezia Gaitskell, Enric Domingo, Bin Li, Tianyang Zhang, Zexi Li, Matthias Friedrich, Dan Woodcock, Clare Verrill, Jens Rittscher

## Abstract

Computational-pathology foundation models (PFMs) have demonstrated remarkable accuracy in a wide range of whole-slide image (WSI) analyses, yet their morphological reasoning and potential biases remain opaque. Here we introduce an attention-shift monitoring framework that tracks tissue-level attention influx and efflux before and after fine-tuning a slide-level aggregator. We apply our interpretable framework across five clinically relevant tasks (lymph-node metastasis detection, lung-cancer subtyping, ovarian-cancer drug-response prediction, colorectal-cancer molecular classification and Marsh grading of colitis). We compare two market-validated PFMs, UNI and prov-GigaPath, using dynamically pooled, compressed embeddings under identical running conditions. Although both models achieve comparable ROC-AUC and balanced-accuracy scores, their attention-shift trajectories diverge sharply: each exhibits broad attention efflux from most tissue regions and highly concentrated, yet minimally overlapping, influx into distinct phenotypic zones. The attention heterogeneity in zero-shot mode and inconsistency of post-tuning attention shifts indicate that the presentation of interpretability depends primarily on each model’s intrinsic feature priors rather than on accuracy or fine-tuning. Our findings uncover a systemic stability gap in PFM interpretability, masked by high performance metrics, and underscore the need for richer explanation tools, bias-monitoring protocols and diversified pre-training strategies to ensure safe clinical deployment.

## Introduction

Artificial intelligence (AI) driven digital pathology has ushered in a new era of histopathological research, enabling unprecedented scale and precision in the analysis of whole slide images (WSIs)^1,2^. As clinical and research laboratories generate ever larger repositories of gigapixel-scale WSIs, deep-learning pipelines have become the de facto approach for tasks ranging from tumour detection to molecular-subtype prediction^3–8^. However, the computational burden of processing these images in their entirety necessitates a multi-stage strategy: first dividing each WSI into fixed-size tiles, then extracting tile-level features, and finally aggregating those features at the slide level^9,10^. In this tiling + feature-modelling paradigm, the fidelity of the tile-level encodings, which encompass biologically meaningful microenvironmental cues and disease-associated bio-semantic representations, is widely recognised as the linchpin for robust downstream performance in computational pathology tasks^11,12^.

Recent advances in large-scale foundation-model pre-training have given rise to Pathology Foundation Models (PFMs), which leverage contrastive learning on massive, and often proprietary, WSI collections to capture rich, fine-grained histological phenotypes^13–17^. Multi-modal variants further integrate textual annotations or genomic profiles, promising a unified representation that spans image and non-image modalities^18–20^. Across a variety of downstream applications, like tumour grading, metastasis detection, survival prediction, PFMs have consistently outperformed traditional handcrafted or shallow-learning feature extractors, underlining their transformative impact on the field^21–23^. Moreover, PFMs assert broad applicability, with their creators often claiming that a single pretrained model can serve the majority of computational pathology and clinical prediction tasks^24–26^.

Notwithstanding these successes, closer examination of PFM development reveals two critical blind spots. First, medical-data privacy and security regulations often restrict public access to training cohorts, rendering the provenance, quality and demographic composition of these WSI repositories opaque^27^, as illustrated in Figure 1a–c, where both tile-level and slide-level data volumes have increased substantially in continuely released PFMs, yet most training cohorts are still derived from limited institutional collaborations. Although some PFMs purportedly draw on multi-centre datasets, essential metadata, such as sampling sites, ethnic and gender distributions, are rarely allowed to be disclosed. When collection locations are reported, they frequently trace back to only one or two local hospitals, raising concerns about single-source bias^15,17,25^. Second, the distribution of cancer types within PFM pretraining sets is markedly imbalanced: in most cases, over half of the images derive from only a handful of cancer types, with long tails of rare or under-represented conditions^15–19,24–26^, as shown in Figure 1d, each PFM is typically dominated by a few major cancer types, while the remaining cancer types contain only a small number of samples. These imbalances threaten to inject systematic biases into downstream model behaviour, particularly in tasks where under-represented phenotypes are clinically salient.

**Figure 1.**
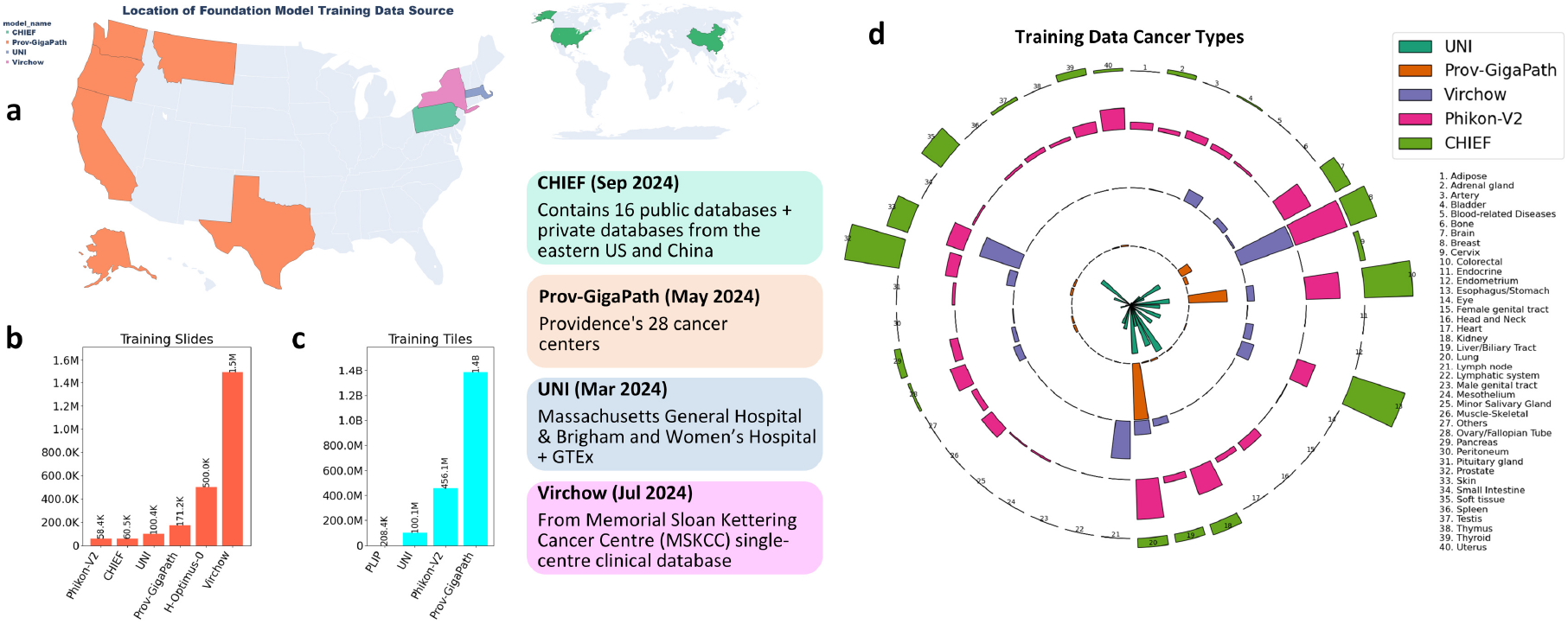
Heterogeneous pre-training contexts for computational pathology foundation models (PFMs). **a**, Geographic sources of training data used by several PFMs. **b**,**c**, Progressive scaling of training data: **b**, at the slide level; **c**, at the tile level. **d**, Imbalanced distribution of cancer types in training datasets across PFMs.

As PFMs are increasingly deployed to support both research and clinical decision-making, it becomes imperative to probe not only their accuracy but also their interpretability and reliability across heterogeneous cohorts and tasks^28^. Yet to date, most PFM evaluations have focused on performance metrics such as ROC-AUC, accuracy and F1 while paying scant attention to how these models arrive at their predictions^21–23,29^. Many PFMs omit fundamental interpretability mechanisms such as attention-based inspection of key tissue regions, concentrating instead on unidimensional accuracy improvements^16,24^. Other approaches that employ slide-level attention visualisations provide only static heatmaps of aggregate scores without elucidating how tile-level feature quality drives attention redistribution and decision-making^15,25,30^. Such simplistic evaluation frameworks and superficial visualisations do not reveal the underlying interpretative processes that govern model behaviour.

Consequently, there is a pressing need for interpretability frameworks capable of elucidating how PFM-derived features guide morphological focus across diverse histological contexts. Here, we introduce an attention shift monitoring framework to address this previously under-explored issue. To ensure the practicality and accessibility of our framework under flexible hardware conditions, we first compress the tile-level features extracted by PFMs, enabling deployment on standard single-GPU environments without compromising downstream performance. We then feed these features into a fixed initialised slide-level aggregator (TANGLE^31^), which is subsequently fine-tuned on specific downstream tasks. By tracking attention influx and efflux at both tile and tissue scales before and after slide-level fine-tuning, our approach captures holistic trajectories of morphological focus and highlights local transition zones in which phenotypic attention fluctuates most dramatically. In doing so, it simulates and monitors the thinking pattern when different PFMs encode and transfer histological knowledge for downstream task-specific reasoning.

Applying this framework to two market-validated PFMs, UNI^15^ and prov-GigaPath^16^, we conduct a comprehensive assessment across five clinically relevant downstream tasks. Notably, the compressed features from both models preserve their original performance, exhibiting negligible impact on accuracy metrics. Furthermore, the two PFMs demonstrate nearly identical predictive performance across all five downstream tasks, thereby offering a controlled basis for evaluating interpretability. These tasks include two widely used public benchmarks, CAMELYON16^32^ (lymph node metastasis detection) and TCGA-Lung^33^ (lung cancer subtype classification), which serve to verify robustness under lightweight computational conditions. Additionally, we evaluate three specialised pathology tasks: Bevacizumab treatment response prediction in ovarian cancer (Ovarian-Bev-Resp)^34^, KRAS molecular subtyping in colorectal cancer (FOCUS-KRAS)^35^, and automatic Marsh grading in colitis (Colitis-Marsh), the last two based on private histopathology cohorts. Despite comparable predictive accuracy, our analysis reveals substantial discrepancies and even opposing attention shift behaviours between the two PFMs, indicating divergent strategies in histological focus redistribution during model adaptation. These findings expose a hidden stability gap in PFM interpretability and underscore the urgency of developing richer explanation tools, robust bias monitoring protocols, and diversified pretraining strategies to ensure safe and reliable clinical deployment.

## Results

### Monitoring knowledge adaptation behaviours of PFMs

We propose a generalisable framework for analysing foundation models’ knowledge transferring behaviour in pathology, with the goal of enabling interpretable deployment of PFMs across diverse histopathological applications. This framework integrates feature compression, task-specific fine-tuning, and multi-perspective interpretative analysis grounded in attention shift tracking (Figure 2). The design follows the widely adopted multiple-instance learning (MIL) paradigm: each whole-slide image (WSI) is tiled into non-overlapping 256 × 256 patches^9^, and each tile is encoded into a feature vector using a pre-trained PFM (Figure 2 a,b).

**Figure 2.**
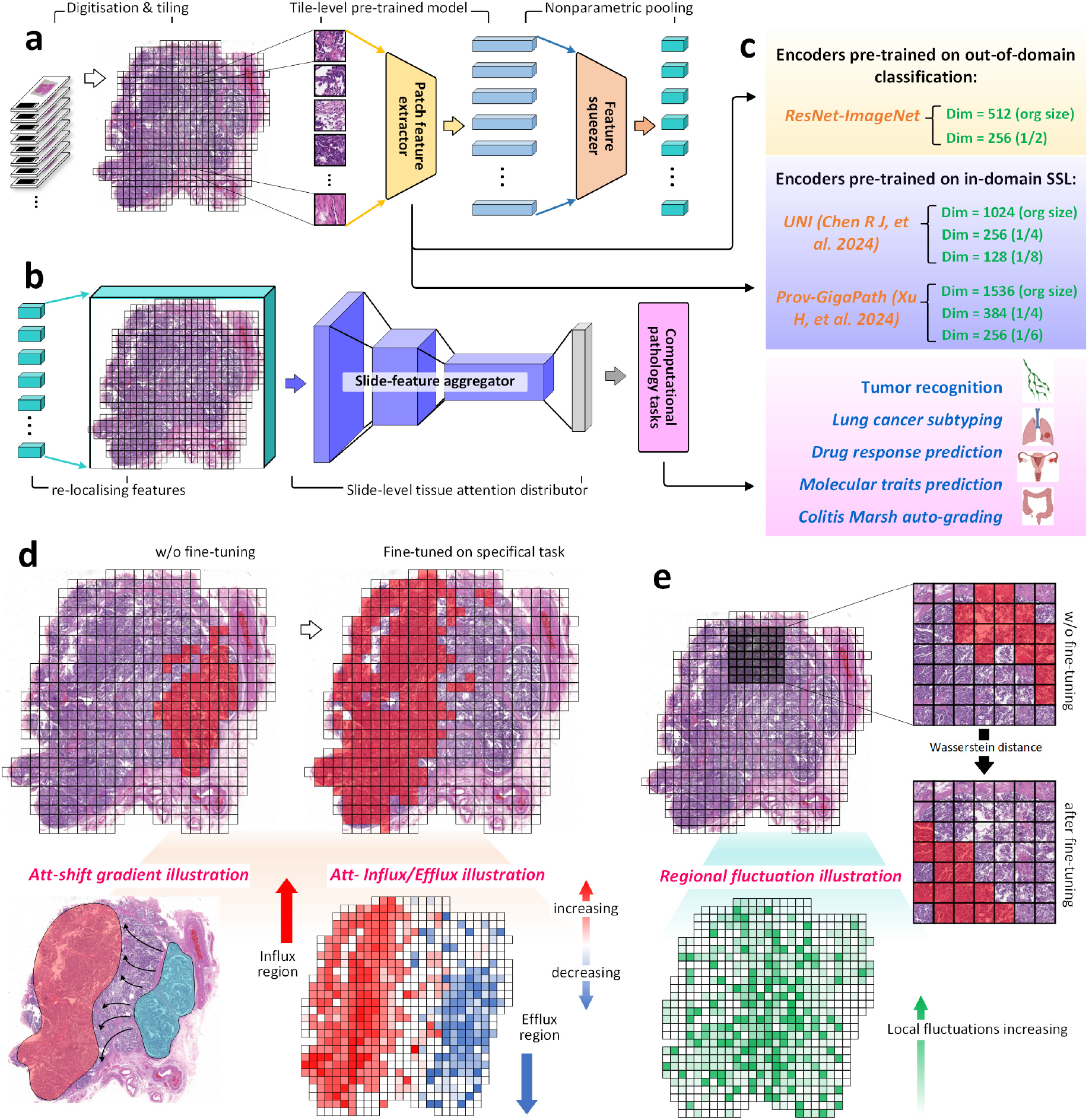
Overview of the framework for fine-tuning on downstream pathology tasks and monitoring attention shift trajectories. **a**, Feature extraction pipeline from whole-slide images, where representations obtained from a foundation model during inference are further compressed through a lightweight module. **b**, The compressed slide-level features are passed into a task-specific aggregator for fine-tuning on downstream objectives. **c**, Comparison of features extracted from ImageNet-pretrained ResNet and two representative pathology foundation models across five diverse downstream tasks. **d**, Monitoring of attention shift trajectories, highlighting the tissue regions where attention flows out and flows into after fine-tuning. **e**, Local attention shift intensity mapping, identifying subregions with high degrees of attention change, which may correspond to transitions in local tissue phenotypic understanding.

To reduce computational overhead without compromising accuracy, we applied a nonparametric pooling mechanism that compresses the high-dimensional tile-level features to lower dimensions. According to our testing, the proposed interpretative framework can be deployed and run stably on a single-GPU environment with as little as 8 GB of memory, making it accessible to almost any research equipment condition. Both compressed and uncompressed features are supported, depending on available computational resources.

The resulting tile representations are passed to a fixed, pre-trained slide-level aggregator, TANGLE^31^, which is subsequently fine-tuned on each downstream task. In addition to generating classification outputs, this aggregator provides spatially resolved attention maps across the whole tissue region, thereby facilitating the interpretation of downstream tasks in line with our analytical objectives.

Our framework is designed to be compatible with a broad range of pathological or typical image foundation models, enabling adaptation and interpretability assessment across diverse feature sources. Specifically, in this study, we evaluated two in-domain PFMs, UNI^15^ and prov-GigaPath^16^, alongside a commonly used out-of-domain baseline, ResNet-18, which was pre-trained on ImageNet^36^. We tested compressed versions of the UNI embeddings at 256 dimensions (1*/*4 of the original size) and 128 dimensions (1*/*8), and prov-GigaPath embeddings at 384 dimensions (1*/*4) and 256 dimensions (1*/*6). ResNet features were compressed from 512 to 256 dimensions. To assess the trade-off between computational efficiency and downstream performance, we also included the original uncompressed representations in all comparisons (Figure 2c).

Our interpretation framework focuses on capturing and visualising the spatial redistribution of model attention before and after slide-level fine-tuning. In particular, it quantifies attention influx and efflux across tissue regions, as well as localised attention fluctuation intensity. These outputs form the basis of our model behavioural analysis and are illustrated in Figure 2(d, e).

To evaluate whether compressed feature representations preserve predictive utility, we benchmarked our framework across five clinically diverse downstream tasks: lymph node metastasis detection (CAMELYON16), lung cancer subtyping (TCGA-Lung), Bevacizumab response prediction in ovarian cancer (Ovarian-Bev-Resp), KRAS molecular subtyping in colorectal cancer (FOCUS-KRAS), and Marsh grading of colitis (Colitis-Marsh). As shown in Figure 3a, both PFMs achieved high ROC-AUC scores across all tasks, with only marginal reductions in accuracy following feature compression. Notably, compressed and uncompressed embeddings from UNI and prov-GigaPath yielded comparable predictive performance across most tasks, and consistently outperformed the ImageNet-trained ResNet-18 baseline. The only exception was in the Colitis-Marsh task, where UNI’s performance was slightly lower than prov-GigaPath (by less than 5%), though the overall improvement over ResNet-18 in this setting was also less pronounced. Balanced accuracy (BACC) results, provided as Figure S1 in the supplementary, support this trend and confirm that feature compression retains essential predictive supportiveness while offering a substantial reduction in computational burden.

**Figure 3.**
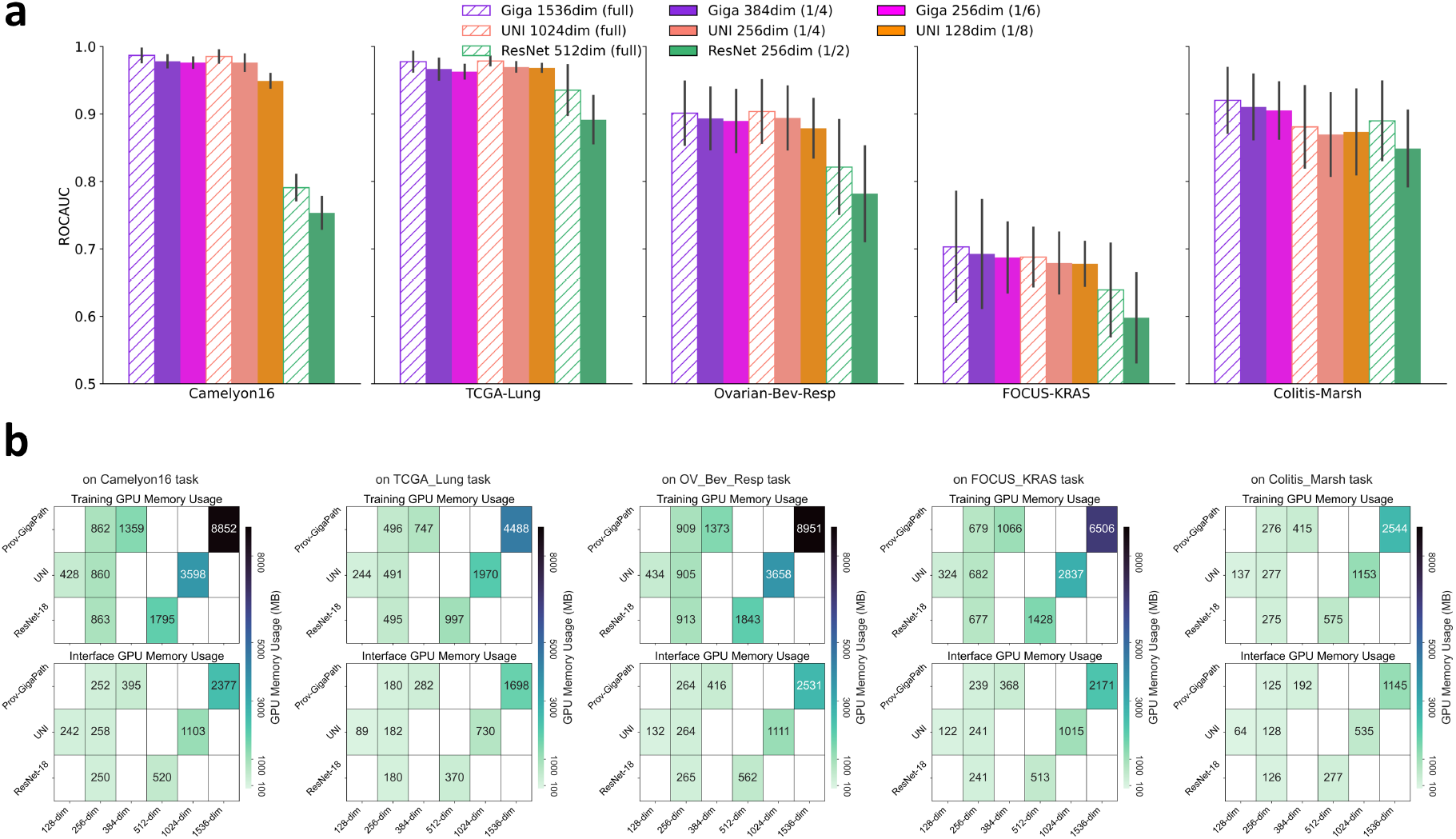
Real-world task performance and memory efficiency of lightweight features. **a**, ROCAUC performance of foundation model features with different compression levels across downstream tasks. Diagonal hatching denotes uncompressed (original) features; solid colours indicate varying degrees of compression. **b**, GPU memory usage for foundation model features of different dimensionalities across tasks.

We also report the corresponding GPU memory usage to estimate the deployability of compressed features across hardware conditions (Figure 3b). Compression enabled up to an eight-fold reduction in memory footprint for UNI and even greater savings for prov-GigaPath, offering practical advantages for large-scale slide processing under constrained computational resources.

We provide a user-facing schematic to illustrate the stepwise application of the proposed interpretation framework across various pathology modelling scenarios (Figure 4). The decision tree begins with a simple distinction: whether the analysis involves foundation models or models trained from scratch. If PFMs are used, the pipeline proceeds through a sequence of mandatory and optional modules, including feature dimensionality compression, slide-level feature aggregation, and task-specific fine-tuning. These steps culminate in the generation of visual outputs, such as attention shift maps and local attention fluctuation profiles. Depending on analytical objectives, users may further apply downstream modules to extract attention influx/efflux patterns, cluster tile-level microenvironmental phenotypes, assess interpretability consistency across different PFMs, and explore cross-task transitions of attention across morphological phenotypes.

**Figure 4.**
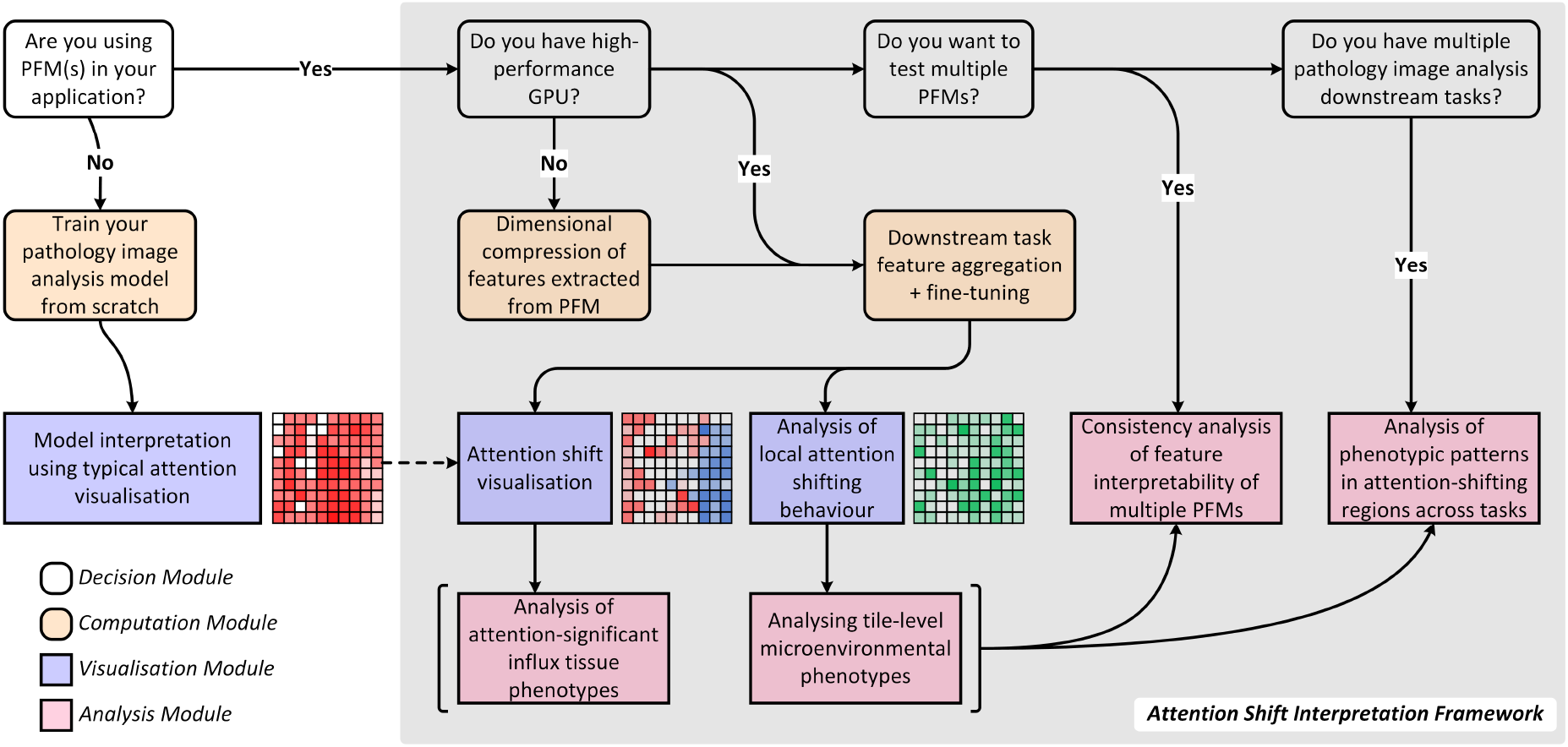
Decision-tree-style user guide for the Attention Shift Interpretation Framework.

Taken together, these modules constitute a broadly applicable and extensible pipeline for monitoring foundation models’ knowledge adaptation behaviours. Beyond enabling performance-preserving compression and efficient fine-tuning, our framework supports systematic assessment of attention redistribution, offering a novel dimension for interpreting model behaviour that complements conventional accuracy-based evaluation and attention-only interpretation. In the following sections, we present our analytical findings step by step, demonstrating how the proposed interpretability framework reveals diverse and sometimes divergent attention behaviours across foundation models.

### Morphological interpretability shaped by PFM embeddings

Pre-trained PFMs capture distinct tile-level histological cues, which we hypothesise shape how morphological focus redistributes during downstream adaptation. We examined this process by tracking attention shifts across tissue regions before and after fine-tuning the slide-level aggregator. For each task, we visualised both large-scale attention migration, characterised by influx into, and efflux out of, specific tissue areas, and localised fluctuation intensity, indicating abrupt transitions in attention focus (Figure 5a–c). Across tasks, we consistently observed that different foundation-model embeddings give rise to markedly divergent attention-shift patterns, revealing distinct morphological reasoning pathways driven by each model.

**Figure 5.**
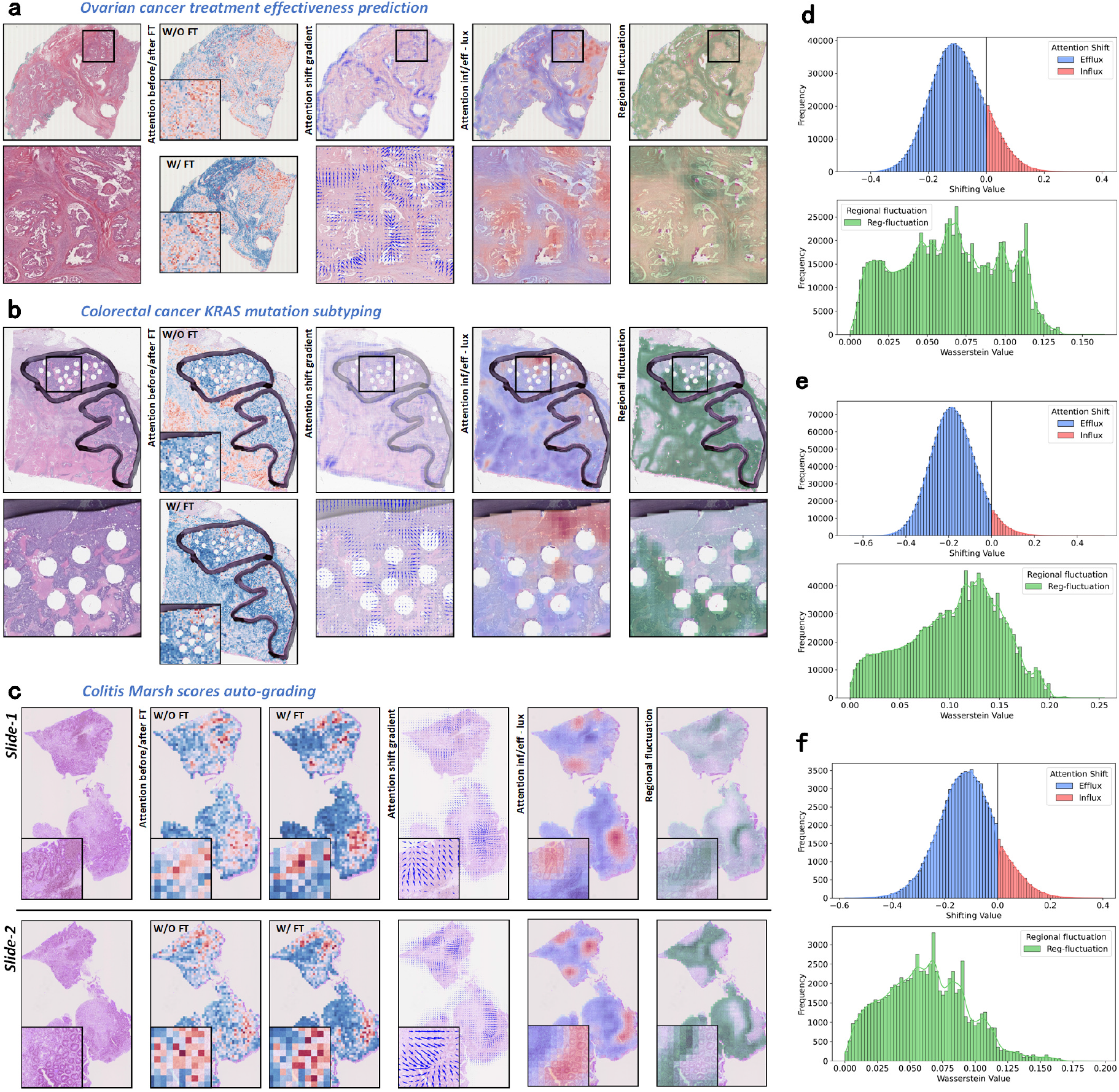
Generation and value distribution of the attention shifting map and local fluctuation map. **a–c**, Visualisation of attention shift and local attention fluctuation across multiple tasks (a: Ovarian-Bev-Resp; b: FOCUS-KRAS; c: Colitis-Marsh). From left to right: original H&E image of an example slide; attention heatmaps before and after aggregator fine-tuning (blue to red indicates increasing attention values from 0 to 1); vector field of attention shift gradients over tissue regions; attention shift heatmap (deeper blue indicates stronger efflux, deeper red indicates stronger influx); local attention fluctuation map (deeper green indicates higher fluctuation intensity). **d–f**, Distribution of attention efflux/influx and local attention fluctuation across slides (d: Ovarian-Bev-Resp; e: FOCUS-KRAS; f: Colitis-Marsh). For each plot, the upper panel shows the distribution of attention efflux (blue bars) and influx (red bars); the lower panel shows the distribution of local attention fluctuation intensity across slides.

While divergent attention-shift patterns were observed across models, certain statistically consistent trends emerged. Notably, attention was broadly withdrawn (efflux) from the majority of tissue regions, with a comparatively smaller subset exhibiting concentrated influx. This general tendency was supported by the distribution curves across multiple datasets (Figure 5d–f), where attention shift values skewed negatively, indicating pervasive efflux behaviour. Upon closer examination, the results in Figure 6d helped to dispel an assumption that attention shifts were primarily determined by initial attention levels, rather than reflecting a task-specific refocus process. Regions exhibiting substantial influx typically did not correspond to those with extremely high or low initial attention. Instead, they were frequently composed of initially moderate-or slightly elevated-attention tiles that were originally considered to bear limited task relevance, but acquired increased task-relevant salience following fine-tuning. Also, the majority of efflux occurred in regions that initially received moderate or lower attention, suggesting we are observing a refinement process of morphological focus rather than a wholesale redistribution.

**Figure 6.**
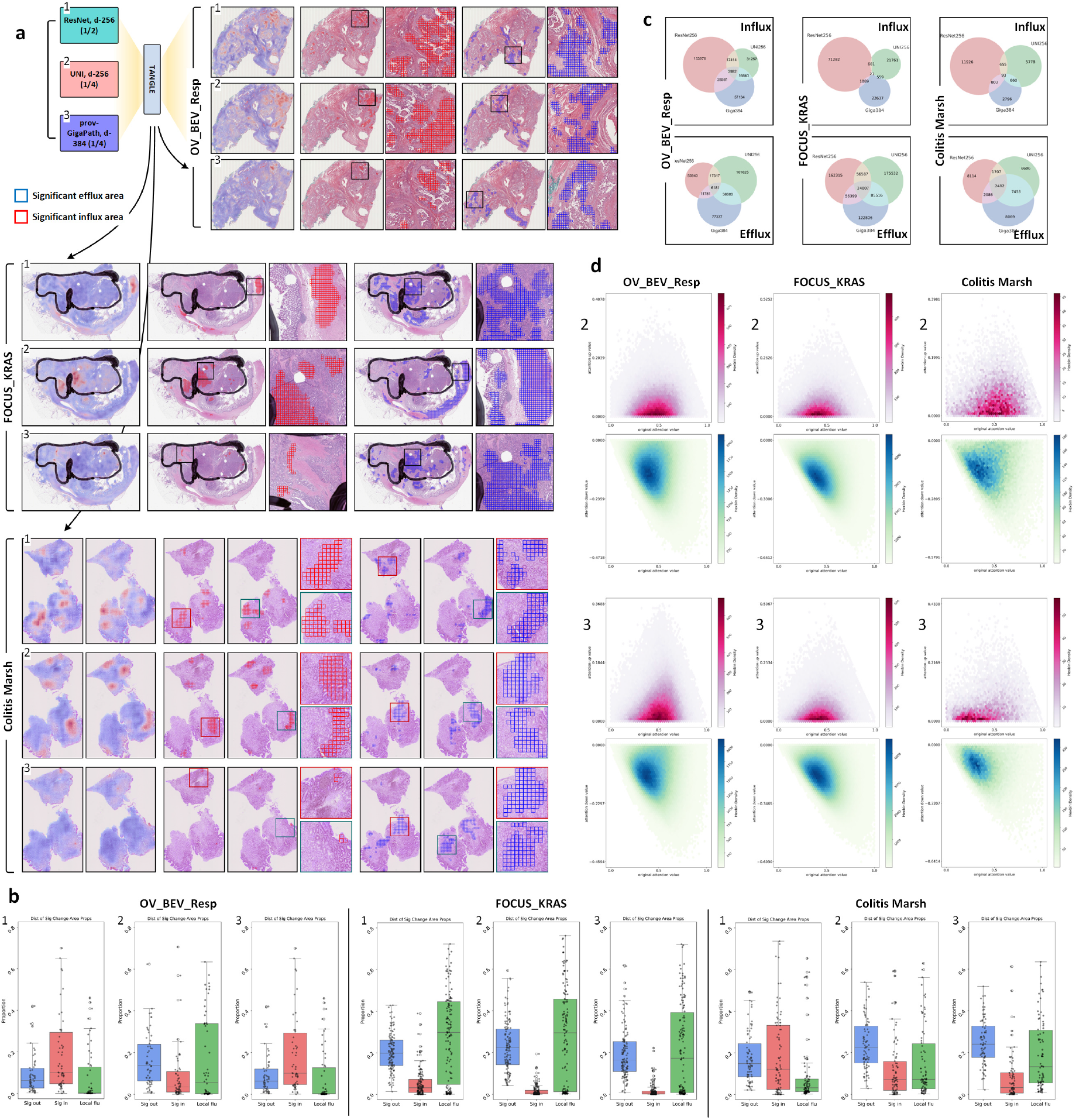
Spatial mapping and distribution of significant attention influx and efflux regions. **a**, Representative tissue regions showing significant attention influx and efflux after aggregator fine-tuning, based on features from three foundation models: ResNet (compressed to 256 dimensions), UNI (256D), and prov-GigaPath (384D). **b**, Proportion of tiles showing significant attention influx, efflux, and strong local attention fluctuation across entire slides after fine-tuning on downstream tasks. **c**, Venn diagrams illustrating the spatial overlap of regions with significant influx and efflux based on different foundation model features after fine-tuning. **d**, Hexagonal binning plots showing the initial attention values and their changes for influx- and efflux-marked tiles, based on UNI (256D) and prov-GigaPath (384D) features.

When we further examined the spatial expression and inter-model concordance of these shifting patterns, representative examples from three downstream tasks visualised the most prominent regions of attention influx and efflux for ResNet-18, UNI, and prov-GigaPath features (Figure 6a), revealing distinct attention influx and efflux landscapes across models. Besides their stronger downstream predictive performance, both PFMs directed their refined attention towards more spatially compact regions that appeared morphologically associated with prediction, such as pathologist-annotated tumour regions or gland-rich areas exhibiting architectural distortion in colorectal tissue, compared with somewhat more diffuse and less biomeanningful focus observed with ResNet-18.

Quantitative analysis of tile-level attention dynamics (Figure 6b) further supported the spatial refocusing patterns described above. Attention influx was concentrated in a relatively small proportion of tiles, typically under 10%, while efflux spanned more extensive tissue areas. Local attention fluctuation, measured via tile-wise Wasserstein distance between pre- and post-fine-tuning attention distributions, was predominantly observed at the peripheries of high influx and efflux regions (see fifth column in Figure 5a–c for visual examples). These fluctuation hotspots suggest pronounced local reconfigurations of attention, which we hypothesise correspond to transitional areas between distinct histological phenotypes, such as tumour-stroma interfaces or epithelial-immune boundaries, where morphological ambiguity is more likely to trigger task-driven interpretative changes.

Nonetheless, comparisons between UNI and prov-GigaPath revealed that, despite exhibiting similar statistical patterns in attention redistribution, the specific regions undergoing significant influx or efflux showed minimal spatial overlap (Figure 6c).

For instance, although both models appeared to direct increased attention towards tumour-associated regions in the ovarian cancer task, the spatial locations and even the associated histological phenotypes varied considerably. In some cases, an influx emerged in nearby yet phenotypically distinct tissue areas. Such divergence reflects the presence of heterogeneous criteria for morphological salience embedded within each PFM’s pre-trained knowledge.

Collectively, these findings indicate that PFMs guide distinct tissue-level morphological interpretations during downstream task adaptation, even when their predictive performance is comparable. The divergence in attention reallocation highlights a key limitation in the stability of interpretability across PFMs, suggesting that even near-identical and highly competitive accuracy may obscure fundamentally different reasoning trajectories between PFMs. These observations motivate a closer investigation, presented in the next subsection, into how such differences manifest across varying downstream tasks and morphological contexts.

### Task-Specific Divergence in Interpretative Behaviour of PFMs

To further investigate how interpretative behaviours are shaped by foundation-model features, we performed cluster-based and spatial analyses of attention influx and local attention shift patterns across three downstream tasks: ovarian cancer treatment response prediction (Ovarian-Bev-Resp), colorectal cancer molecular subtyping (FOCUS-KRAS), and colitis severity grading (Colitis-Marsh). For each task, we identified tile-level attention influx clusters and visualised their spatial organisation on tissues, associating each cluster with its underlying histomorphological phenotype (Figures 7–9a). In parallel, we examined high-frequency local attention shift patterns, focusing on localised transitions across phenotypically distinct regions (Figures 7–9b).

**Figure 7.**
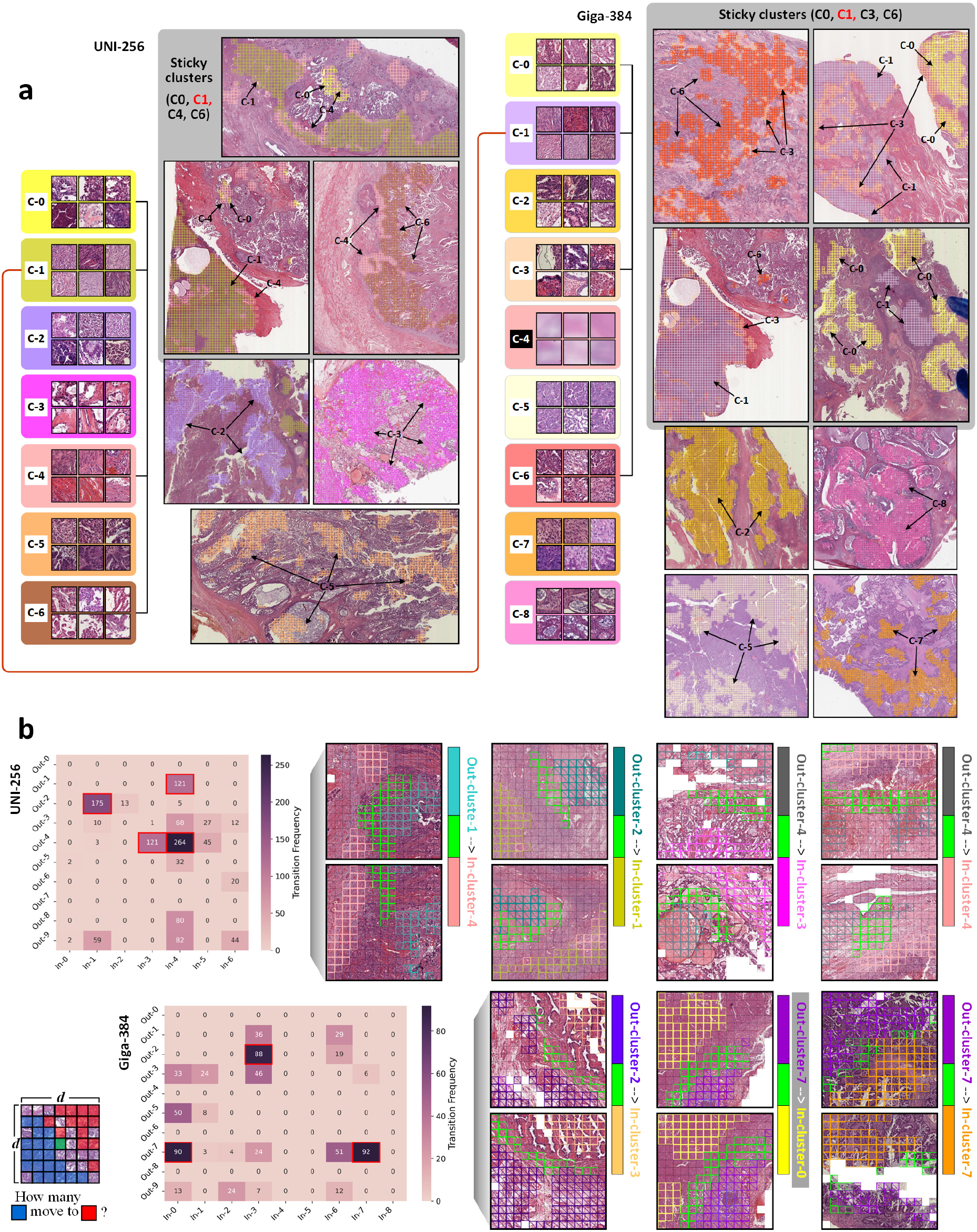
Heterogeneous attention shifting behaviours in the ovarian cancer treatment response prediction task. **a**, Morphological phenotypes of attention influx regions identified using Gaussian Mixture Model clustering based on features from UNI (256D) and prov-GigaPath (384D). Representative phenotypes of each cluster are shown (left and right panels), along with their spatial distributions in tissue. Clusters highlighted with a grey background indicate spatial adjacency on tissue. Red lines connect cluster pairs with substantial spatial overlap, as further illustrated in Supplementary Materials. **b**, Analysis of local attention shift behaviours surrounding high-fluctuation regions. Heatmaps display transition frequencies from efflux clusters to influx clusters (with high-frequency transitions highlighted in red boxes), and example tissue regions illustrating spatially coherent transition pathways.

**Figure 8.**
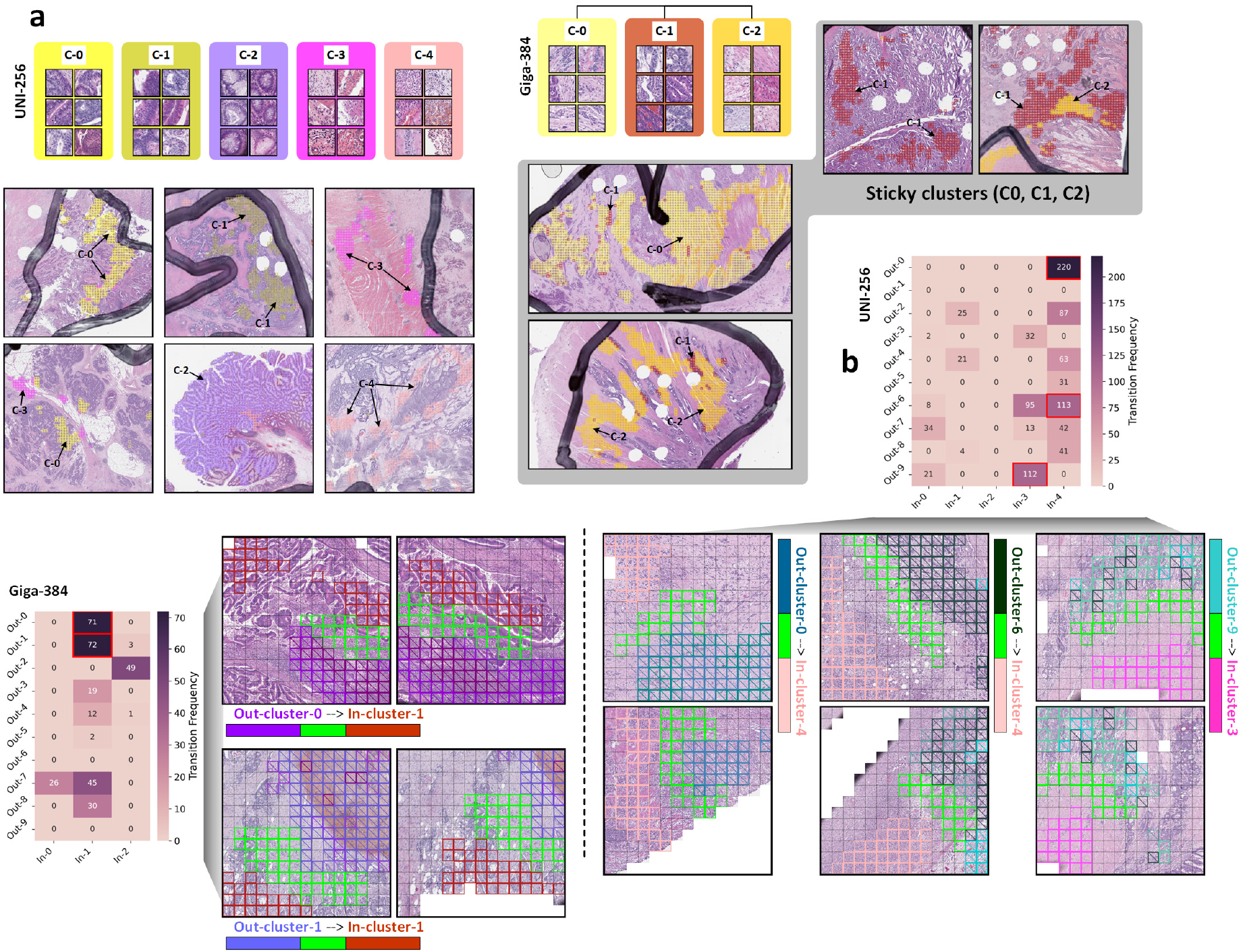
Attention shift behaviours in the colorectal cancer molecular subtyping task. **a** and **b**, Visualisation and clustering results based on the FOCUS-KRAS dataset, using features from UNI and prov-GigaPath. Subfigure structure and interpretation are consistent with Figure 7.

**Figure 9.**
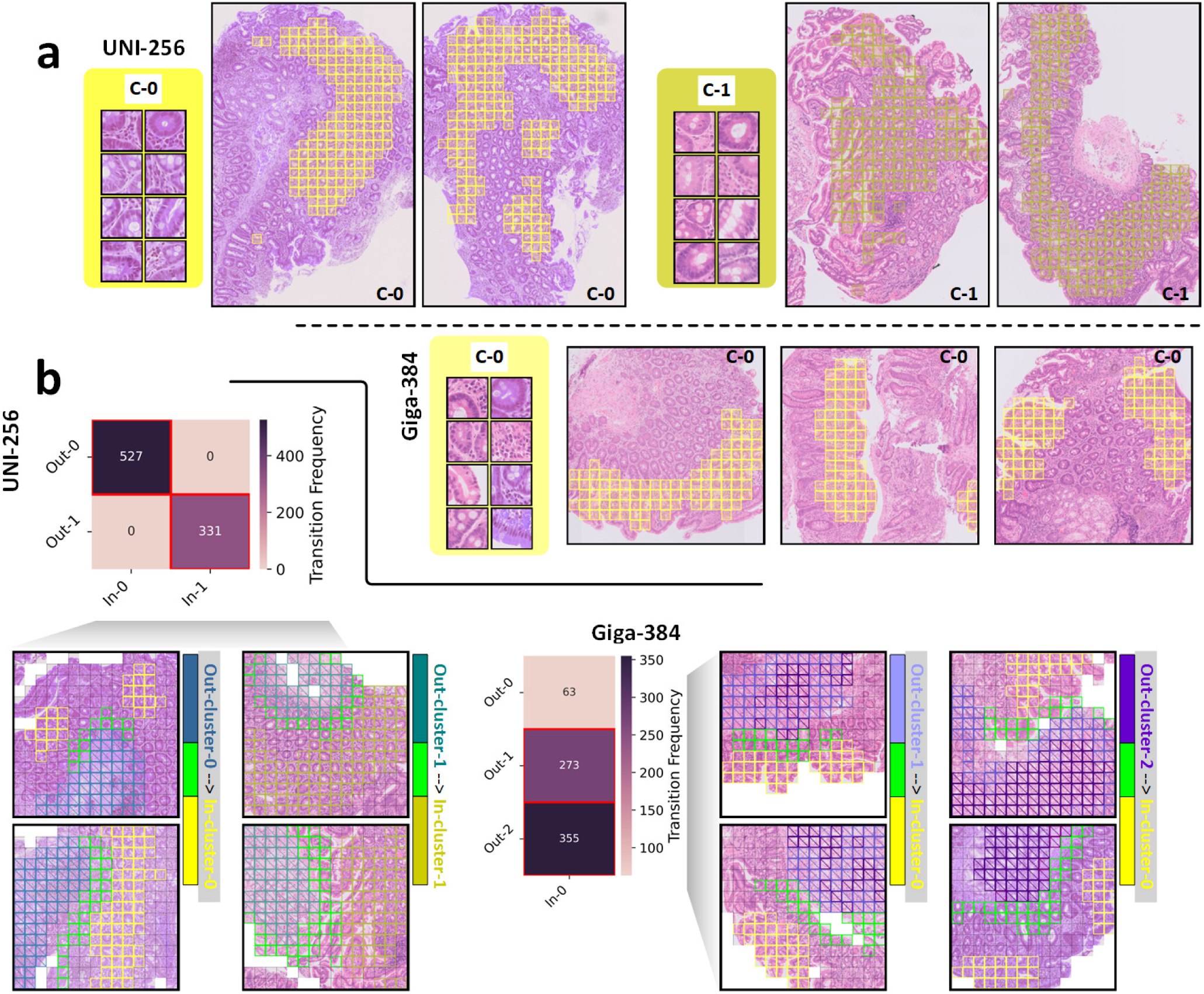
Attention shift behaviours in the colitis Marsh auto-grading task. **a** and **b**, Visualisation and clustering results based on the Colitis-Marsh dataset, using features from UNI and prov-GigaPath. Subfigure structure and interpretation are consistent with Figure 7 and 8.

For each task, we found that the tiles exhibiting significant attention influx could be consistently grouped into a limited number of clusters, automatically determined without manual supervision. These clusters, obtained from different PFMs, each displayed distinct histological features, except cluster C-4 derived from GigaPath features in the Ovarian-Bev-Resp task, which corresponded to morphologically uninformative areas and was excluded from further investigation. Importantly, the attention influx clusters were not spatially scattered but instead occupied spatially contiguous regions on tissue, indicating that they each represented coherent morphological zones rather than isolated features without bio-organisation.

Although all PFMs produced spatially compact clusters of attention influx, the specific locations and associated histological phenotypes of these clusters were rarely concordant across models. Except for a single partially overlapping case in the ovarian cancer task, attention influx clusters derived from UNI and GigaPath exhibited largely distinct spatial footprints and histomorphological characteristics (also shown in Supplementary Figure S2). For example, in the colorectal molecular subtyping task, UNI frequently attended to tumour epithelial zones characterised by densely packed acinar structures and proliferative activity, while GigaPath instead prioritised adjacent fibrotic stromal areas with smooth muscle–like spindle cell components, both morphologically structured, yet divergent in microenvironmental composition and phenotypic implications. Such divergence suggests that each PFM identifies and prioritises task-relevant morphological cues in its own unique and internal understanding, thereby constructing model-specific interpretative pathways while performing the same downstream task.

In some cases, multiple attention influx clusters identified by the same PFM were found to occupy spatially adjacent or continuous tissue regions, despite representing distinct histological features. This observation suggests that PFM-derived features can direct attention across morphologically diverse but spatially coherent areas, potentially reflecting interpretative focus within histologically heterogeneous tissue compartments. However, even in these scenarios of local continuity, the spatial co-occurrence between influx clusters derived from different PFMs remained minimal.

Further evidence of internal interpretative consistency within each model came from the distribution of attention influx clusters across predictive classes (Supplementary Figure S4). Each PFM exhibited a non-uniform class association across its clusters, with most of the clusters more frequently appearing in one predictive class than the others. This indicates that attention reallocation was not arbitrary, but aligned with each model’s internal decision logic, producing class-specific patterns of interpretative focus. In addition, most attention influx and efflux clusters were not restricted to specific cases but were distributed broadly across slides within each dataset (Supplementary Figure S5), suggesting that the attention shifting signals extracted by each PFM generalise across diverse tissue samples.

We further examined the green-highlighted tiles (in Figures 7–9b), which mark hotspots of sharp attention fluctuations within their peripheral regions. These areas reveal fine-grained attention shift behaviours across transitional tissue contexts, offering insights into how PFMs modulate localised attention patterns at histomorphological boundaries. Notably, in downstream tasks, we frequently observed contrasting local attention trajectories across PFMs. For instance, in the FOCUS dataset, UNI-based embeddings shifted attention from reactive stromal regions towards adjacent tumour-dense areas, whereas GigaPath embeddings exhibited the opposite trend, focusing instead on extracellular matrix (ECM)-rich regions. These divergent patterns suggest that even within the same local tissue context, PFMs may adopt distinct interpretative strategies when encountering morphologically ambiguous or transitional structures.

Taken together, this analysis illustrates that the diversity in interpretative behaviour across PFMs is not confined to high-level summary statistics, but also manifests in both global and local spatial attention shifting trajectories. Each PFM constructs a distinct and internally coherent interpretive paradigm, shaped by its pretraining context and representational biases. These paradigms give rise to task-specific and markedly divergent reasoning pathways across models. These findings motivate further investigation into the generalisability of such interpretive divergence across different datasets.

### Task-Agnostic Interpretative Behaviour Divergence across PFMs

To assess whether the distinct interpretative behaviours driven by different PFMs extend beyond task-specific contexts, we conducted a cross-task clustering analysis of tile-level attention influx and efflux features. Specifically, we aggregated all tiles exhibiting significant attention shifts across the five downstream tasks and respectively clustered them using a unified, model-agnostic visual feature space. These clusters capture broadly recurring histological phenotypes that underlie meaningful reallocation of attention across diverse pathological scenarios. To evaluate whether PFMs exhibit consistent or divergent interpretative behaviour at this cross-task circumstance, we performed bidirectional cross-assignment tests: each cluster corresponding to one PFM (e.g., UNI) was projected into the joint influx & efflux feature space of the other PFM (e.g., GigaPath) to examine alignment in attention shift directionality.

Results from this cross-task analysis confirmed the generalisability of each PFM’s attention-shifting patterns. As shown in Supplementary Figure S6, histological phenotype clusters with significant attention influx and efflux were not restricted to individual tasks but instead appeared across multiple datasets. Although the distribution of these phenotypes varied by cohort, both PFMs demonstrated specific task-agnostic attention redistribution patterns.

However, comparison between UNI and GigaPath revealed substantial divergence in their task-agnostic attention behaviours. As illustrated in Figure 10 along with Supplementary Figure S6, only two pairs of clusters demonstrated directional agreement in attention assignment across PFMs, whereas five pairs exhibited mismatches, e.g., a cluster associated with attention influx in one model corresponded to efflux in the other. These mismatches suggest that the same histological phenotype could be differentially interpreted, or even inversely weighted, depending on the model’s pretraining knowledge and the internal salience criteria it enacts. Taken together, these findings extend the PFM-specific nature of interpretability beyond individual tasks, revealing that foundational divergence in attention logic persists across pathological domains and manifests in globally distinct reasoning preferences.

**Figure 10.**
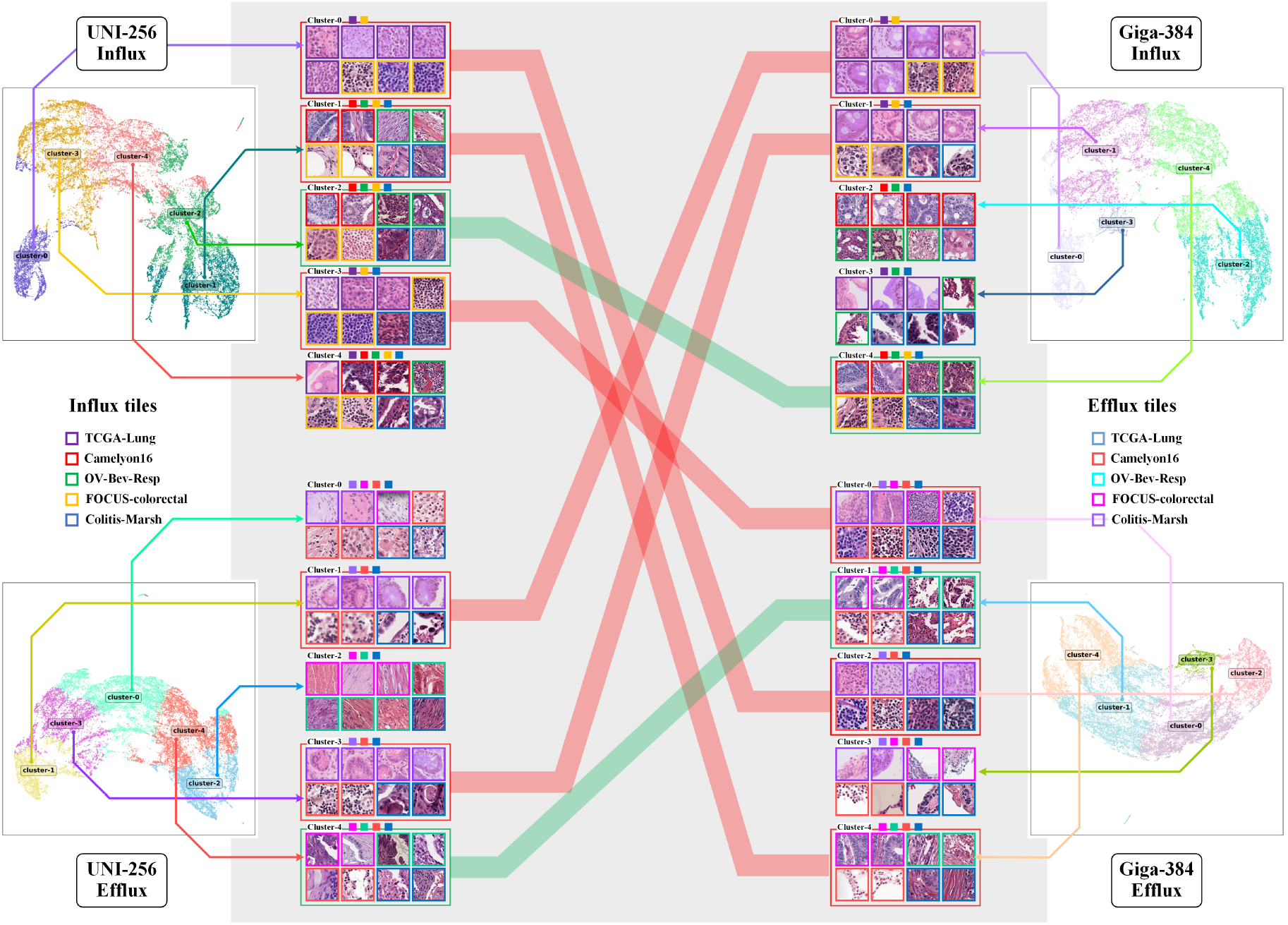
Task-agnostic heterogeneity of interpretable evidence. Cross-task clustering of histological phenotypes associated with significant attention influx and efflux, based on features from UNI (256D) and prov-GigaPath (384D). Each side shows UMAP visualisations and representative tile examples for the identified clusters; tile border colours indicate the downstream task dataset of origin. Bars in the centre connect cluster pairs with high spatial alignment across the two feature sets. Green bars denote agreement in attention direction (both influx or both efflux), while red bars indicate mismatches (influx in one, efflux in the other).

## Discussion

### Multi-scale Interpretation of Attention Shifts

In this study, we proposed an interpretable framework for monitoring the knowledge adaptation behaviours of pathology foundation models (PFMs), with a particular emphasis on spatial attention redistribution during downstream task execution. Our framework incorporates two complementary components, attention-shift trajectories and local fluctuation maps, to uncover how pretraining priors influence morphological focus during downstream task adaptation. While attention-shift maps reveal large-scale changes in attention reallocation across tissue structures, local fluctuation maps highlight phenotypically transitional areas where attention reconfiguration is especially pronounced. Together, these elements facilitate a multi-scale interpretation of model behaviour, beyond conventional attention heatmaps.

### Tile-Level Feature Determines Morphological Interpretation

A straightforward observation in our study is the decisive influence of tile-level feature quality on downstream interpretability. Under a controlled setting with fixed aggregator architecture and identical fine-tuning protocols, we found that tile-level features extracted from different PFMs guided different attention distributions at both the zero-shot and post-fine-tuning stages. The initial, zero-shot attention states varied across models, reflecting an unstable foundational understanding of task-relevant histomorphology. Furthermore, the magnitude and direction of subsequent attention shifts were not merely a function of initial allocation, but were more strongly driven by each PFM’s intrinsic representation of pathological phenotype. These results reveal that PFMs may converge in predictive accuracy, yet diverge substantially in interpretative behaviour, a discrepancy that would remain invisible under traditional performance metrics.

A classic example of differences shaped by pretraining backgrounds is technical artefacts in tissue preparation, such as sectioning angle, which can alter the apparent morphology (e.g., crypts appearing circular in central regions versus finger-like at the tissue perimeter). Such artefacts, along with the overall quality of pretraining data, may introduce biases in PFMs at the level of visual feature representation, thereby influencing how models prioritise histological regions. These feature-level biases also provide a partial explanation for the persistent interpretative divergence observed across different PFMs.

### Persistent Interpretative Divergence Among PFMs

Notably, the observed interpretability heterogeneity persisted across task-specific and task-agnostic settings. In individual downstream cohorts, attention-influx clusters derived from different PFMs exhibited little spatial or phenotypic overlap, even when producing similar predictive outcomes for the same classification task. Furthermore, local attention transitions near morphologically heterogeneous regions exhibited model-specific directionalities, which in some cases were reversed across PFMs. When clustering attention-shifting tiles across all downstream datasets, we found that even task-agnostic attention redistribution behaviour remained markedly divergent, where identical histological phenotypes were often associated with attention influx in one model but efflux in the other. These findings collectively underscore that PFM reasoning trajectories are not just diverse, but are shaped by deeply embedded priors that fine-tuning cannot easily override. Such divergences do not directly demonstrate erroneous behaviour, as exemplified by the Colitis-Marsh task, where different PFMs prioritised distinct yet still relevant epithelial structures for grading (e.g., UNI emphasising centrally located crypts, and prov-GigaPath highlighting border-adjacent epithelium, as illustrated in Figure 11). Nevertheless, the lack of convergence remains a concern for the stability of interpretability and underscores the need for cautious, task-aware scrutiny of attention-shifting patterns.

**Figure 11.**
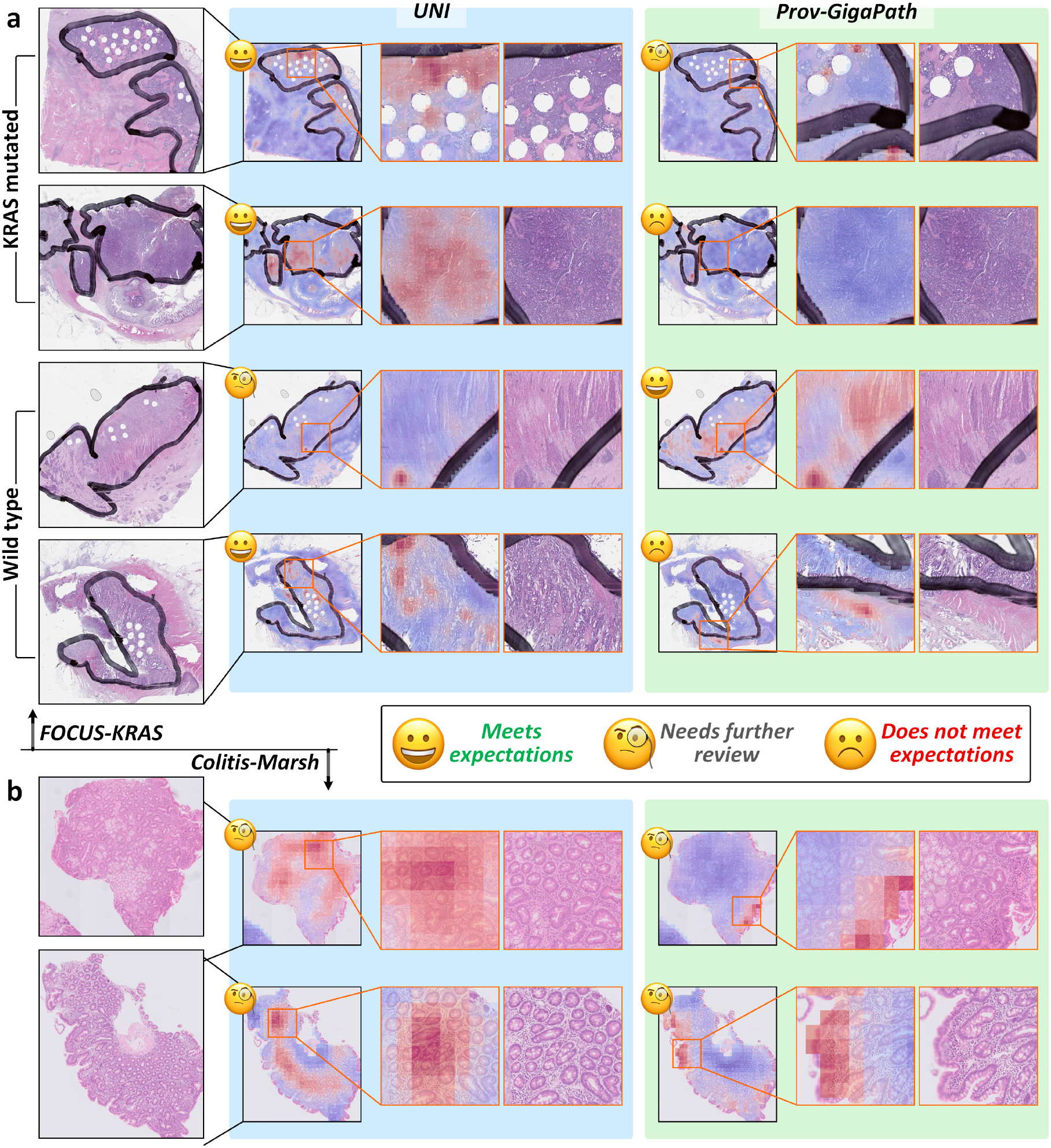
Expert review of divergent attention-shifting behaviours in the FOCUS-KRAS and Colitis-Marsh datasets. Attention-shift visualisations (red: attention influx, blue: efflux) were generated using features from UNI and prov-GigaPath, and experts evaluated these visualisations; their assessments were summarised into three interpretative levels: *Meets expectations, Needs further review*, and *Does not meet expectations*. **a**, KRAS-mutated and wild-type colorectal cancer samples, where UNI frequently redirected attention to invasive adenocarcinoma with glandular differentiation, whereas prov-GigaPath favoured adenoma-like or early-stage carcinoma regions, at times displaying opposite attention flows. **b**, Colitis-Marsh cases, where UNI tended to focus on central tissue regions, while prov-GigaPath more often highlighted the tissue borders.

These divergences highlight not only the instability of interpretability across PFMs but also the importance of providing practical guidance for how such interpretability assessments can be meaningfully employed by domain experts in real-world tasks.

### Practical use cases for the interpretability framework

Beyond investigating PFM stability, our framework also offers constructive guidance for how domain experts may employ interpretability assessments in practice. Across datasets, pathologists noted that attention-shifting behaviours were generally consistent within each model, yet diverged sharply between PFMs, underscoring that these signals are systematic rather than random. Based on this, we envisage the following application scenarios:

- In settings where clinicians have deep familiarity with private cohorts and specific tasks, expert review of attention influx and efflux can be used to judge whether the focus of a PFM is biologically plausible. For example, in the ovarian drug-response task, shifting attention away from tumour cores towards peritumoural stroma may still be interpretable in light of treatment-associated microenvironmental changes. By contrast, in the FOCUS-KRAS task, experts expected attention to remain concentrated in invasive tumour regions with marked atypia and desmoplastic stroma. As shown in Figure 11, UNI aligned with this expectation, while prov-GigaPath often redirected focus to glandular differentiation patterns with more elongated tubular or villous structures, occasionally even reversing UNI’s attention pattern. Although prov-GigaPath achieved slightly higher accuracy, such behaviour highlights the possibility that improved performance metrics may obscure biologically less plausible reasoning pathways.
- Automation can be increased by integrating locally deployed large language models (LLMs)^37^ to generate histomorphological descriptions of attention influx and efflux regions. Pathologists found that these automatically produced narratives were of sufficient quality to enhance convenience and accelerate review, facilitating comparison of systematic differences in model behaviour. Examples of such histomorphology-based descriptions of attention influx and efflux patterns at different scales, generated by the locally deployed LLM, are provided in the Supplementary and shown as Figure S7–S12.
- Multimodal indicators may provide an external reference point for adjudicating whether attention shifts are clinically meaningful. For instance, one could examine whether attention redistribution correlates with proteomic or genomic features, or whether it associates with prognostic measures such as survival outcomes. Such triangulation could further validate whether observed interpretative focus reflects robust biological signals or spurious patterns.

The aforementioned use cases illustrate how the proposed framework can be operationalised to help pathologists and clinicians probe foundation models in a task-specific manner, detect potential misinterpretation risks, and safeguard against overfitting masked by aggregate accuracy. At the same time, it is important to note that PFMs may economise on representational capacity and reasoning effort by selecting only a subset of informative features, rather than all equally relevant structures. Examining attention shifts can help to detect if the model learned features that come from batch effects (e.g. scanners/staining protocols in different centres). This also underscores the need to maintain a cautious attitude when interpreting attention maps and related interpretability outputs, while also motivating the development of more comprehensive interpretability tools.

### Systemic Challenges for Foundation Model Interpretability

Although our experiments focused on two widely used pathology foundation models, the broader implications likely extend across the current PFM landscape. Most existing PFMs rely on similar training paradigms and do not introduce fundamentally novel mechanisms for histomorphological reasoning or interpretability. Thus, the distinct behaviours revealed by our framework reflect not incidental differences, but structural limitations in the stability of model reasoning under current pretraining practices. Our work thus calls for more critical scrutiny of PFM interpretability and suggests that performance parity across models should not be equated with consistency in biological insight.

### Translational Impact and Prospects for Safe Deployment

The proposed interpretability framework provides a practical means for evaluating pathology foundation models (PFMs) before their application in diagnostic or investigative settings. Acting as an interpretability safeguard, it allows domain experts to examine whether a model’s reasoning behaviour is consistent with established pathological understanding by monitoring attention-shift patterns across on specific cohort. This process helps to determine whether the model’s prior knowledge and adaptation dynamics are appropriate and biologically coherent for the task at hand.

At the same time, highly consistent yet unexpected patterns of attention redistribution observed across diverse PFMs may serve as potential indicators of previously unrecognised biological phenomena, suggesting opportunities for further hypothesis generation and discovery. The framework fulfils a dual role, functioning both as a safeguard for interpretability and as a perspective for exploring novel biological insights.

The framework also offers a reference for how future PFMs can be evaluated and refined. Developers may use attention-shift monitoring as part of pre-release safety assessment, testing models across heterogeneous datasets to ensure that their interpretative behaviours remain stable and biologically plausible.

### The Need for More Comprehensive Interpretability Frameworks

Our findings also highlight an urgent demand for more expressive and trustworthy interpretability frameworks. Standard attention maps, while simple and convenient, offer limited granularity and operate on fixed representational states, failing to capture the nuanced ways in which pretraining priors embedded within PFMs shape attention behaviour in morphologically complex regions. By contrast, our framework enables both global and local interpretative assessments grounded in tissue morphology, revealing PFMs’ distinct behavioural patterns previously hidden from view. As foundation models become increasingly central in computational pathology, we advocate that more interpretability tools, along with infrastructures for evaluating their reliability, must co-evolve to ensure safe, transparent, and fair deployment of such models.

### Challenges and Prospects

Finally, our results raise critical questions about the origins and mitigation of pretraining bias in PFMs. Given the privacy constraints and fragmented nature of medical data, driving scaling laws in pathological pretraining remains a formidable challenge. We advocate for the development of robust bias monitoring strategies and knowledge-balancing mechanisms during the construction of PFMs^38,39^. Promising directions may involve multi-centre pretraining under federated learning^40^ frameworks, as well as model fusion schemes^41^ that integrate complementary knowledge from diverse training sources. Although an ideal scenario would involve different PFMs converging on similar attention-shift behaviour, which could be taken as an optimal situation of interpretability robustness. In practice, a lack of convergence to a similar interpretability pattern does not necessarily invalidate a model’s utility, as different PFMs may still rely on distinct but biologically relevant features to achieve comparable accuracy. Rather, such divergence highlights the need for cautious interpretation and indicates that more prospective trials can ultimately establish the true predictive value of these models as clinical biomarkers.

## Methods

### Ethics statement

Ethical approval of the Ovarian-Bev-Resp dataset was obtained under protocols TSGHIRB No.1-107-05-171 and No.B202005070, and all data were anonymised prior to retrospective analysis^34^. The FOCUS cohort was reviewed and approved as part of S:CORT consortium by the South Cambs Research Ethics Committee (REC ref 15/EE/0241).

### Data Acquisition and Preprocessing

This study utilised five histopathological datasets spanning both public benchmarks and proprietary clinical cohorts. Three datasets, CAMELYON16^32^, TCGA-Lung^33^, and Ovarian-Bev-Resp^34^, were sourced from public repositories and cover lymph node metastasis detection, lung cancer subtyping, and ovarian cancer treatment response prediction, respectively. Two additional datasets, FOCUS-KRAS^7,35^and Colitis-Marsh, were curated from institutional archives to support evaluation in KRAS molecular sub-typing in colorectal cancer and autoimmune disease grading. Details of each dataset are summarised below.

#### CAMELYON16

The CAMELYON16 dataset comprises 399 whole-slide images (WSIs) of haematoxylin and eosin (H&E)-stained sentinel lymph node sections, collected from two medical centres in the Netherlands: Radboud University Medical Centre (Nijmegen) and University Medical Centre Utrecht. The cohort includes 270 slides for training and 129 for testing, with a total of 239 normal and 160 metastasis-labelled slides. Slides were scanned at 40×, 20×, and 10× magnifications. The dataset was originally developed to benchmark automated metastasis detection methods in breast cancer patients and is widely adopted in computational pathology research.

#### TCGA-Lung

The TCGA-Lung dataset consists of 1,053 diagnostic WSIs drawn from two subtypes of lung cancer: lung adenocarcinoma (LUAD) and lung squamous cell carcinoma (LUSC). The images were acquired from multiple institutions worldwide as part of routine clinical care under the TCGA framework, contributing to substantial heterogeneity in scanning protocols and device manufacturers. We discarded one corrupted slide and randomly partitioned the remaining 1,053 slides into 842 for training and 211 for testing. All slides were downsampled to 20× and 5× magnifications. Only slide-level diagnostic labels were available for this dataset.

#### Ovarian-Bev-Resp

This private dataset comprises 288 H&E-stained WSIs from 78 patients with epithelial ovarian cancer, sourced from the Tri-Service General Hospital and the National Defence Medical Centre, Taipei, Taiwan. Slides were digitised using a Leica AT2 scanner at 20× magnification, with an average image resolution of 54,342 × 41,048 pixels (27.34 × 20.66 mm). Each case is labelled based on clinical response to Bevacizumab therapy, with 162 treatment-effective and 126 non-effective cases. Associated clinical metadata includes serum CA-125 levels, histological subtype, and treatment outcome. **FOCUS-KRAS**. The FOCUS-KRAS dataset was derived from the FOCUS clinical trial conducted in the UK, which compared therapeutic strategies in advanced colorectal cancer. A total of 666 H&E-stained WSIs from 362 patients were initially collected, all scanned at 20× magnification using Aperio systems. After removing the unlabelled samples, 658 slides were retained for analysis. Tumour regions were annotated by expert gastrointestinal pathologists, with rigorous exclusion of necrotic and artefactual tissue. This cohort was developed under the S:CORT consortium and is paired with rich genomic profiling, including RNA microarrays and targeted DNA sequencing. Pathological and molecular annotation protocols followed standardised quality controls for downstream analysis. Population characteristics of the external validation subset used in this study are summarised in Supplementary Table S1.

#### Colitis-Marsh

The Colitis-Marsh dataset consists of H&E-stained slides from patients with colitis, annotated for Marsh grading of mucosal inflammation severity. The dataset comprises 304 tissue samples collected from 67 patients, with Marsh grade labels distributed as follows: grade-0: 80, grade-2: 43, grade-3: 66, grade-4: 80, and grade-5: 35.

#### Preprocessing of Whole-Slide Images

All WSIs were processed in a consistent pipeline to ensure quality and comparability across datasets. Each slide was first read using the `OpenSlide`^1^ library, an open-source toolkit for reading virtual slides in multiple formats. Preprocessing involved the removal of irrelevant or artefactual regions, including: (1) grey or white background; (2) areas contaminated by blue or green staining artefacts; and (3) ink marks from black, red, green, or blue annotation pens. These operations were implemented using `OpenCV`^2^ image processing tools.

After cleaning, tissue regions were tiled into fixed-size patches. For CAMELYON16, tiles of 256× 256 pixels were extracted at 40× magnification (0.25, *µm* per pixel). For all other datasets, the same tile size was used but extracted at 20× magnification (0.5, *µm* per pixel). Only tiles with at least 70% tissue content, as determined by pixel-level thresholding, were retained for downstream processing.

### Tile-Level Feature Extraction and Compression

Let each WSI be divided into a set of non-overlapping tiles 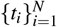, where each tile *t*_*i*_ is represented as a fixed-size RGB image of shape *H* ×*W* × 3. To encode the morphological information within each tile, we define a feature extractor *ℱ*_*θ*_: ℝ^*H*×*W* ×3^ → ℝ^*d*^, where *θ* denotes the pre-trained parameters of the PFM and *d* is the original feature dimensionality. This yields a set of tile-level feature vectors 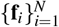, where **f**_*i*_ = *ℱ*_*θ*_ (*t*_*i*_) and **f**_*i*_ ∈ ℝ^*d*^.

To facilitate deployment of the proposed framework in typical clinical or institutional settings, where access to high-performance computing clusters may be limited, we apply a non-parametric compression function 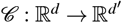 that maps the original feature space to a lower-dimensional latent space, where *d*′ *< d*. The compression is implemented via a dynamic average pooling mechanism, which divides the input vector into *d*′ non-overlapping consecutive segments of equal length, then averages each segment to produce a single output dimension.

Formally, for a feature vector **f** ∈ ℝ ^*d*^, the compressed vector 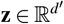 is computed as:

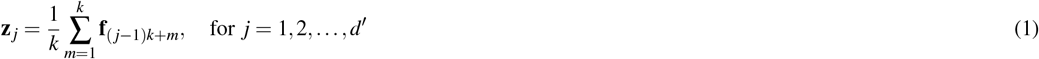

where **z**_*j*_ is the *j*-th element of the compressed feature vector **z**, obtained by averaging over a contiguous segment of 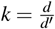 dimensions from the original feature **f** ∈ ℝ^*d*^.

In our experiments, we extracted tile-level features from two publicly available PFMs: UNI^15^ and prov-GigaPath^16^, both pretrained using contrastive learning on large-scale WSI repositories. To establish a baseline, we also used ResNet-18 pretrained on ImageNet as an out-of-domain comparator. The original feature dimensionality was *d* = 1024 for UNI, *d* = 1536 for prov-GigaPath, and *d* = 512 for ResNet. These were compressed as follows:

- UNI: compressed to *d*′ = 256 and *d*′ = 128, representing 1/4 and 1/8 of the original size;
- prov-GigaPath: compressed to *d*′ = 384 and *d*′ = 256, representing 1/4 and 1/6 of the original size;
- ResNet: compressed to *d*′ = 256, i.e. 1/2 of the original size.

This compression strategy enables our interpretability framework to operate under limited hardware resources without seriously compromising predictive fidelity.

### Slide-Level Aggregator and Fine-Tuning

To obtain slide-level predictions from the extracted tile-level features, we employed the TANGLE^31^ with attention-based multiple instance learning (ABMIL)^42^ structure as the aggregator. This module was pre-trained on the TCGA-BRCA cohort and thus initialised with fixed parameters capturing slide-level priors for invasive breast cancer. For all downstream tasks in this study, we retained this pretraining and fine-tuned the aggregator parameters, while keeping all tile-level features frozen.

Given a slide be represented as a set of *n* tile embeddings {**f**_1_, **f**_2_, …, **f**_*n*_}, where **f**_*i*_ ∈ℝ^*d*^. The aggregator computes a weighted representation of the slide by learning attention scores over tiles through a gated mechanism. Each attention vector **a**_*i*_ is defined as the element-wise product of two transformation branches:

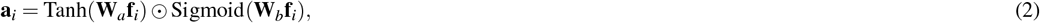

where 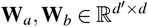 are learnable projection matrices, and ⊙ denotes the Hadamard product. These attention vectors are projected onto a learnable vector 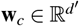 to compute normalised weights using a softmax function:

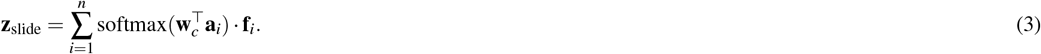

The resulting slide-level representation **z**_slide_ is subsequently passed to a linear classifier to produce the final task-specific prediction. During downstream task adaptation, only the aggregator parameters were updated; tile-level features from the PFMs were kept frozen.

All downstream fine-tuning procedures were conducted under identical optimisation settings to ensure comparability across models and tasks. A learning rate of 1 × 10^−4^ was used with a batch size of 4. The optimiser employed was Adam with weight decay set to 1× 10^−3^, accompanied by an exponential learning rate scheduler with decay factor *γ* = 0.95. Each model was trained for a fixed number of 20 epochs without early stopping.

### Evaluation of Prediction Tasks

We employed two evaluation metrics to assess the predictive performance of each foundation model in downstream tasks: the area under the receiver operating characteristic curve (ROC-AUC) and balanced accuracy (BACC). These metrics were chosen to reflect both discrimination capacity and robustness under class imbalance.

#### ROC-AUC

This metric quantifies the model’s ability to distinguish between positive and negative classes across a range of classification thresholds. Formally, it is defined as the probability that a randomly chosen positive instance is assigned a higher score than a randomly chosen negative instance:

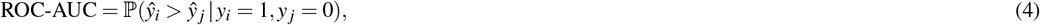

where *ŷ*_*i*_ and *ŷ*_*j*_ are the predicted scores for positive and negative samples, respectively, and *y*_*i*_, *y* _*j*_ ∈ {0, 1} are their corresponding ground truth labels.

#### Balanced Accuracy

This provides an unbiased estimate of classification performance under class imbalance by averaging the recall obtained on each class. For binary classification, it is computed as:

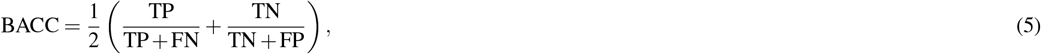

where TP, TN, FP, and FN denote true positives, true negatives, false positives, and false negatives, respectively.

For each downstream task, we report both ROC-AUC and BACC based on slide-level predictions derived from the aggregator outputs. To ensure robust and fair comparisons between models, we employed cross-validation protocols stratified by class distribution. Specifically, for datasets with relatively limited sample sizes (Ovarian-Bev-Resp and Colitis-Marsh), we performed 10-fold cross-validation. For the remaining tasks (CAMELYON16, TCGA-Lung, and FOCUS-KRAS), we used 5-fold cross-validation. In all settings, slides were randomly split into training and testing partitions with an 80%/20% ratio in each fold. The reported performance metrics represent the mean values across all folds for each task.

### Attention Shift and Local Fluctuation Map Generation

We visualised model focus at multiple scales using four complementary map types: (i) static slide-level attention maps at a given stage (before or after fine-tuning), (ii) an attention-shift map that quantifies influx/efflux between stages, (iii) a block-wise gradient vector field that summarises movement directions, and (iv) a local fluctuation map that highlights sharp reconfiguration of attention within small neighbourhoods. Static attention visualisation follows the standard attention-based MIL practice used in ABMIL^42^, CLAM^9^, and TANGLE^31^; the latter three maps are derived from pre-/post-fine-tuning attention states.

#### Notation

For a slide tiled into *n* tiles, let 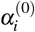 and 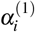 denote the scalar attention assigned to tile *i* before and after fine-tuning, respectively (Subsection: Slide-Level Aggregator and Fine-Tuning). Let *𝒢* be the 2D tile grid. We obtain tile-grid attention maps *𝒜* ^(*s*)^: *𝒢* → ℝ_≥0_ by placing 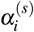 at the spatial cell of tile *i*, with *s* ∈ {0, 1}. A binary tissue mask *ℳ*: *𝒢* → {0, 1} suppresses background. To overlay on the slide thumbnail, grid maps are upsampled to image coordinates via bilinear interpolation (`OpenCV`) and rescaled by min–max normalisation, denoted Norm_[*a,b*]_(·), which maps values into a fixed range [*a, b*] over tissue pixels.

#### Static attention maps

The static attention map at stage *s* is simply the normalised, masked map

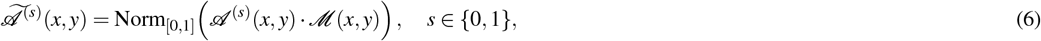

which corresponds to the conventional attention heatmap used in TANGLE^31^.

#### Attention-shift map (influx/efflux)

To quantify redistribution after fine-tuning, we compute the signed difference between post- and pre-fine-tuning maps, followed by local smoothing and range normalisation. Let

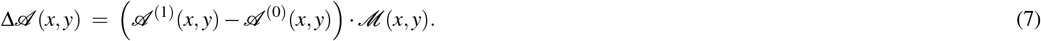

For each grid location (*x, y*), let *N*_*k*_(*x, y*) denote its *k* ×*k* neighbourhood (with reflect padding) and | *N*_*k*_(*x, y*) | its size. A locally smoothed map is obtained as

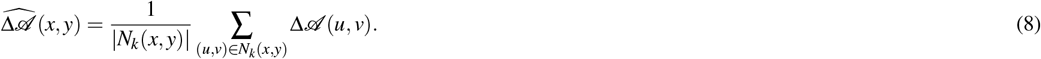

The result is rescaled to [−1, 1] and [0, 1] for visualisation:

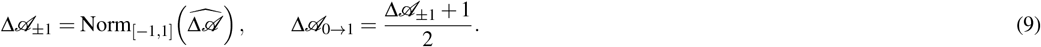

Positive values indicate attention influx and negative values indicate efflux. In practice, we used *k* = 5 unless stated otherwise.

#### Block-wise gradient vector field of attention shift

To summarise spatial movement directions, we compute a coarse vector field from the smoothed shift map. For a block size *b*× *b*, let ⟨ · ⟩_*b*_ denote block averaging over non-overlapping *b* ×*b* regions. Finite-difference gradients of the block-averaged map yield

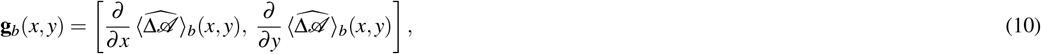

and only vectors with ∥**g**_*b*_(*x, y*) ∥ ≥*τ* are retained, plotted as a quiver field over the slide thumbnail (coordinates scaled to thumbnail size). We used *b* = 7 and *τ* = 0.1 by default.

#### Local fluctuation map via Wasserstein distance

Local reconfiguration of attention is captured by comparing within-window distributions before and after fine-tuning. For each pixel (*x, y*), let Ω_*w*_(*x, y*) denote the *w* × *w* neighbourhood on the tile grid and |Ω_*w*_| its cardinality. Define two empirical 1D distributions formed by the attention values in that window,

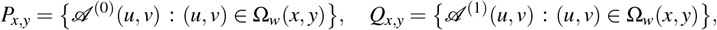

with uniform weights on samples. The Wasserstein-1 distance (Earth Mover’s Distance^43^) is

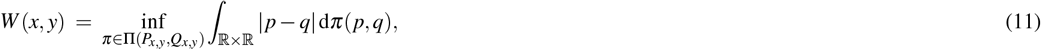

where Π(*P, Q*) denotes the set of couplings with marginals *P, Q*. To discount uniform regional drifts (e.g., when all values move coherently up or down), we compute a correction factor from the locally averaged raw shift:

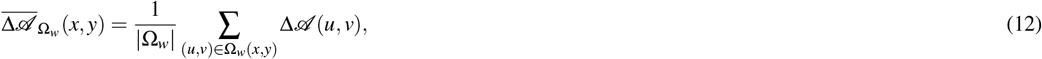

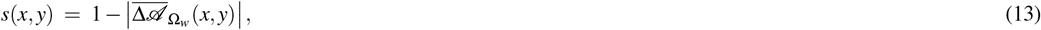

and define the filtered fluctuation score

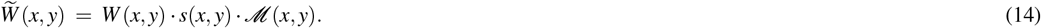

Both *W* and 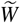 are finally normalised to [0, 1] over non-zero tissue pixels (sparse min–max normalisation). Unless otherwise specified, we used *w* = 7 and reflect padding.

#### Parameterisation of maps

All maps are computed at the tile grid resolution and upsampled to the slide thumbnail. As mentioned above, the attention-shift map uses a 5 × 5 uniform filter; the gradient field uses block size 7 × 7 (tiles) and magnitude threshold *τ*=0.1; the local fluctuation map uses window size 7 × 7 (tiles) with drift correction as in Eq. (13). Background and pen-mark regions are excluded via the tissue mask *ℳ* (Subsection: Data Acquisition and Preprocessing). Positive/negative codes for influx/efflux follow Eq. (9).

### Macro-Level Statistical Analyses

Before performing dataset-specific analyses of spatial phenotypes, we conducted a set of macro-level statistical procedures to quantify global patterns of attention redistribution across PFMs.

### Selection of significant tiles

For each slide in the held-out test sets, we identified tiles exhibiting the strongest changes in attention values between pre- and post-fine-tuning states. Specifically, the top 5% of tiles with the largest positive shifts (influx) and the bottom 5% with the largest negative shifts (efflux) were extracted from the attention-shift map (Subsection: Attention Shift and Local Fluctuation Map Generation). These tiles, together with their immediate neighbourhoods, were retained for subsequent analyses.

### Proportion of significantly affected tiles

For each slide, the fraction of significant tiles was computed relative to the total number of valid tissue tiles:

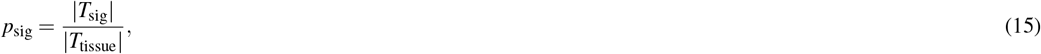

where *T*_sig_ denotes the set of significant influx, efflux, or locally fluctuating tiles, and *T*_tissue_ the full set of tissue-containing tiles. These per-slide proportions were aggregated within each cohort.

#### Cross-model overlap of significant tiles

To assess cross-model concordance, we compared the sets of significant tiles across PFMs. For a given slide and shift direction (influx or efflux), let *U*_*m*_ denote the set of significant tiles for model *m* ∈ {ResNet-18, UNI, prov-GigaPath}. Pairwise and three-way intersections between {*U*_*m*_} were computed to quantify overlap:

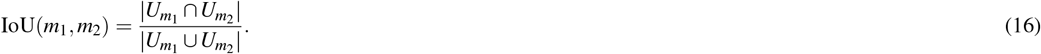

Venn diagrams were used for visualisation of the joint distributions of *U*_*m*_ for influx and efflux tiles separately.

#### Correlation with baseline attention

Finally, we examined the relation between initial attention values and subsequent changes after aggregator fine-tuning. For each tile *i*, we defined the pair

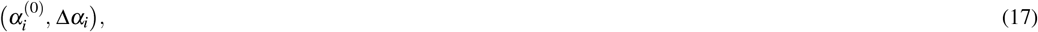

where 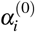 is the pre-fine-tuning attention weight and 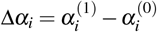 the change after fine-tuning. Separate sets of pairs were collected for influx and efflux tiles. Their joint distributions were summarised via hexagonal binning density estimation, which provides robust aggregation over large numbers of tiles and accounts for the non-Gaussian nature of attention changes.

### Spatial Pattern Analysis

In the approach of interrogating the spatial organisation of attention redistribution, we analysed slide-level patterns in three downstream tasks: Ovarian-Bev-Resp, FOCUS-KRAS, and Colitis-Marsh. Each analysis comprised two modules.

#### Morphological clustering of influx tiles

For each PFM, we collected all tiles within the held-out test sets that exhibited significant attention influx (top 5%, cf. Subsection: Macro-Level Statistical Analyses). Each tile was represented by its PFM-derived feature embedding. To investigate the morphological phenotypic subtypes indicated by attentional influx behaviours driven by different PFMs, we performed unsupervised clustering using Gaussian Mixture Models (GMMs)^44^, which was implemented using the `scikit-learn` library^3^. A GMM models the feature distribution as a weighted sum of multivariate Gaussians:

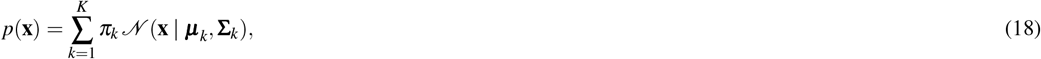

where *π*_*k*_ are the mixture weights (∑_*k*_ *π*_*k*_ = 1), and ***µ***_*k*_, **Σ**_*k*_ denote the mean and covariance of component *k*. The optimal number of components *K* was selected by minimising the Bayesian Information Criterion (BIC):

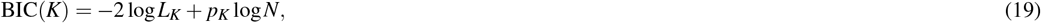

where *L*_*K*_ is the maximised likelihood under a *K*-component GMM, *p*_*K*_ is the number of free parameters, and *N* is the number of tiles. We restricted *K* ≤ 10 to avoid over-fragmentation. Clusters corresponding to artefacts or background contamination were excluded. The retained clusters were subsequently examined for their spatial configuration within slides, their adjacency relationships, and the degree of overlap between clusters identified by different PFMs.

#### Local co-occurrence of influx and efflux

In parallel, we investigated whether strong attention influx and efflux co-occurred within the same local neighbourhoods. For each slide, we first computed local fluctuation maps via Wasserstein-based metrics (cf. Subsection) to identify regions of sharp redistribution. Within these regions, we enumerated the frequency with which tiles from influx clusters and efflux clusters were observed in spatial proximity. By aggregating statistics across slides, we obtained a transition frequency matrix indexed by out-clusters (efflux) and in-clusters (influx). This matrix was visualised as a heatmap, with high-frequency entries highlighted, thereby pinpointing local spatial motifs where attention shifting flows were consistently activated. Top-occurring transitions were further traced back to representative slides for qualitative inspection.

### Cross-Cohort Analyses

When we interrogate whether attention redistribution patterns generalise across cohorts, we conducted cross-dataset analyses that jointly considered all test slides from Ovarian-Bev-Resp, FOCUS-KRAS, and Colitis-Marsh.

#### Feature representation

To ensure fairness independent of PFM-specific embeddings, we represented tiles using hand-crafted descriptors based on Local Binary Patterns (LBP). For a given pixel at position (*x, y*) with grey-level value *I*(*x, y*), the LBP code is defined as

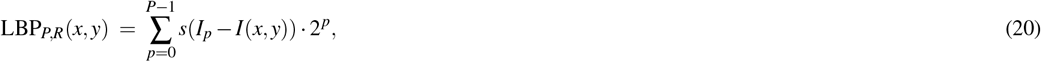

where *P* is the number of sampling points evenly spaced on a circle of radius *R*. The neighbour intensities *I*_*p*_ and the thresholding function *s*(*z*) are defined as

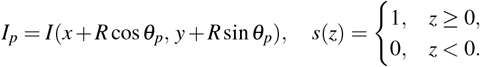

Here *θ*_*p*_ = 2*πp/P*, and *I*_*p*_ is obtained via bilinear interpolation at the non-integer coordinates. The resulting LBP codes are aggregated into a histogram with (*P* + 2) bins under the ‘uniform’ variant^45^. Following standard practice, we set *P*=24 and *R*=3, which provides a balance between discriminative power and computational cost. We computed such histograms independently on each RGB channel and concatenated them, yielding a 78-dimensional feature vector per tile. This provided a common visual feature space that is invariant to the backbone used for attention assignment.

#### Clustering procedure

Within this feature space, we separately aggregated tiles showing significant influx and efflux (cf. Subsection: Macro-Level Statistical Analyses) for UNI and prov-GigaPath. Each of the four groups (UNI-influx, UNI-efflux, prov-GigaPath-influx, prov-GigaPath-efflux) was clustered using *k*-means clustering^46^ with *k*=5, thereby producing five canonical phenotypic clusters per group.

#### Cross-model projection

To assess concordance between PFMs, we projected clusters derived from one model into the joint influx/efflux cluster space of the other. Concretely, each tile cluster identified from UNI was reassigned to the nearest cluster centroid computed from prov-GigaPath (and vice versa), based on Euclidean distance in the LBP feature space. We then recorded whether the reassigned cluster label preserved the original influx/efflux designation. This allowed us to quantify to what extent the redistribution phenotypes detected by one PFM were reproducible in the other, and whether systematic mismatches (e.g. influx mapped to efflux) occurred.

### LLM-based Generation of Histomorphological Descriptions

We used large language models (LLMs) to produce concise, pathology-oriented descriptions of regions showing significant attention redistribution for complementing visual and statistical analyses. Two pipelines were employed.

#### (A) Describing representative visualisations with a general-purpose vision–language LLM^47^

We used *GPT-5*^4^ to generate free-text reports for a curated set of visualisations from the test public dataset - Ovarian-Bev-Resp and each PFM (UNI and prov-GigaPath). For this investigated dataset, three types of inputs were prepared: (i) randomly sampled tile examples from clusters with significant attention influx; (ii) tissue thumbnails with coloured grids indicating the spatial location of influx clusters on tissue; and (iii) illustrations of high-frequency regional transitions from efflux to influx. Each interaction began with a short role-setting prompt, followed by a task-specific question. Table 1 lists the exact prompts. Placeholders such as `<specific colour>` were programmatically replaced with the colour names used in the highlight areas of visualisation.

**Table 1.** Prompts used with Llama-3.2-11B-Vision for representative visualisations.

| Dataset / Input type | Initial prompt | Question prompt |
| --- | --- | --- |
| Ovarian-Bev-Resp / Tiles from influx clusters | You are a pathologist and need to review some image analysis results. I will continue to provide you with a set of 256 x 256 pathology images. These are H&E-stained patched biopsies of ovarian cancer patients. You will be asked to help review each set of images and describe the histological phenotypes they exhibit. | Could you tell me what histological phenotypic features are presented in these patches as a whole? |
| Ovarian-Bev-Resp / Spatial context of influx clusters | You are a pathologist, and you need to help me review some pathology image analysis results. These images are H&E-stained pathology images from patients with ovarian cancer. I will now show you images of sections of a large tissue, some of which are marked with coloured grids. You need to focus on the areas marked with specific colours and describe the morphological phenotypes they reveal. | Tell me what morphological phenotype the exactly <specific colour> grid indicated area shows? |
| Ovarian-Bev-Resp / Regional efflux→influx transitions | You are a pathologist, and you need to help me review some pathology image analysis results. These images are H&E-stained pathology images from patients with ovarian cancer. I will now give you some pathological images within a local tissue range, in which boxes of different colours indicate different small areas. I will ask you about the tissue histomorphological phenotypes exhibited by 1-2 of the colour-indicated areas. | What histomorphological phenotypes are present in the regions indicated by <colour-a> and <colour-b>? What phenotypic evolution does the region undergo? |
| FOCUS-KRAS (all three input types) | Same as above but “colorectal cancer patients”. | Same as in Ovarian-Bev-Resp. |
| Colitis-Marsh (all three input types) | Same as above but “colitis patients”. | Same as in Ovarian-Bev-Resp. |

#### (B) Batch descriptions for all tiles in influx/efflux clusters with a medical-specific LLM^48^

We also used locally deployed LLM - *LLaVA-Med v1*.*5 (Mistral-7B)*^5^ to generate short descriptions for every tile belonging to clusters with significant attention influx or efflux (for both UNI and prov-GigaPath). For each tile, we queried the model with a three-question set tailored to the dataset; prompts are shown in Table 2. The three answers were concatenated to form a single per-tile narrative.

**Table 2.** Prompts used with LLaVA-Med v1.5 for tile-wise batch description.

| Dataset | Questions (asked sequentially for each tile) |
| --- | --- |
| Ovarian-Bev-Resp | 1) How to describe the pathological phenotypes in this image of H&E-stained ovarian cancer tissue?<br>2) Is there any anomalies described in the micro-environment of the given image?<br>3) What types of cells and biological features are shown in the area shown in the given image? |
| FOCUS-KRAS | 1) How to describe the pathological phenotypes in this image of H&E-stained colorectal tissue?<br>2) Is there any anomalies described in the micro-environment of the given image?<br>3) What types of cells and biological features are shown in the area shown in the given image? |
| Colitis-Marsh | 1) How to describe the pathological phenotypes in this image of H&E-stained colon tissue from colitis patients?<br>2) Is there any anomalies described in the micro-environment of the given image?<br>3) What types of cells and biological features are shown in the area shown in the given image? |

#### Post-processing and visualization

During the word cloud and term frequency (TF) analysis in pipeline (B), the common English stop-words were removed, and punctuation and numerical tokens were discarded. The calculation of word cloud and TF statistics separately for UNI and prov-GigaPath in their influx versus efflux clusters. Word-clouds (shown in Supplementary Materials) were rendered with the `wordcloud` package^6^ using the TF weights, and bar charts display the most frequent terms (the same preprocessing was used across datasets to ensure comparability).

### Expert Interviews on PFM Interpretability

In order to complement the quantitative analyses, we conducted semi-structured interviews with domain experts in pathology, oncology and biology. The purpose was to evaluate the proposed attention-shift monitoring framework in practice and to collect professional insights on the biological plausibility of attention-shifting behaviours exhibited by different PFMs.

On three downstream datasets (Ovarian-Bev-Resp, FOCUS-KRAS, and Colitis-Marsh), attention-shift visualisations derived from both UNI and prov-GigaPath were prepared. For each diagnostic, prognostic or molecular subtype class within a dataset, we randomly selected 3 - 5 representative cases, resulting in a set of paired visualisations that contrasted the two PFMs. These materials were reviewed independently by clinical experts with domain expertise specific to the corresponding cohort (e.g., gynaecological oncology, gastrointestinal pathology, or inflammatory bowel disease).

Each interview lasted approximately 40 - 60 minutes and was conducted in an open, conversational format. No rigid questionnaire was imposed, but discussions were guided around three broad themes: (i) the extent to which experts would place trust in the outputs of pathology foundation models, and what forms of interpretability evidence could strengthen such trust; (ii) how experts perceived the attention-shift behaviours shown by different PFMs, including whether these were considered biologically meaningful and which outputs aligned more closely with their expectations; and (iii) the perceived role of attention-based interpretability in future pathology workflows, including how manual or automated validation could help establish the reliability of model-derived evidence.

The expert feedback was qualitatively analysed and incorporated into the Discussion section, where it is used to contextualise interpretability divergences across PFMs and to propose practical use cases for the framework. While the interviews were exploratory in nature, they provided constructive insights that grounded the interpretability analyses in real-world clinical reasoning.

### Computational Environment and Software Dependencies

All experiments were implemented in `Python`-3.12 and `PyTorch` v2.3.1 with `CUDA` 12.1 support. Inference and fine-tuning were performed on a server equipped with an Intel(R) Xeon(R) Gold 6242 CPU (32 cores, 2.80 GHz), 376 GB system memory, and four NVIDIA RTX 6000 GPUs (24 GB memory each); however, each model was fine-tuned/tested using a single GPU only. All fine-tuning and inference routines were designed to fit within an 8 GB GPU memory budget, ensuring the feasibility of deployment on economical research workstations.

Whole-slide images (WSIs) were accessed and processed using the `OpenSlide` library. Foundation model weights were managed and loaded through the `Hugging Face Transformers`^7^ APIs. Image preprocessing employed `OpenCV`, while clustering analyses were performed with `Scikit-learn`. Visualisations were primarily generated with `Matplotlib`^8^ and extended using higher-level libraries such as `Plotly`^9^, `Seaborn`^10^, and `matplotlib-venn`^11^.

All source code used in this study will be made publicly available at https://github.com/superhy/placeholder upon publication.

## Supporting information

Supplementary dataset information and results

## Data Availability

All data produced in the present study are available upon reasonable request to the authors
All data produced are available online at public repositories or requested from the authors

## Funding

CV and JR were supported by the PathLAKE Centre of Excellence for digital pathology and artificial intelligence, which is funded by the Data to Early Diagnosis and Precision Medicine strand of the HM Government’s Industrial Strategy Challenge Fund, managed and delivered by Innovate UK on behalf of UK Research and Innovation (UKRI) (Grant ref: 104689/application number 18181). YH and JR were supported by Novo Nordisk A/S - Big Data Institute (BDI), University of Oxford collaboration project. CV was supported by the NIHR Oxford Biomedical Research Centre. GB was supported by Fergus Gleeson’s A2 research funds, UKRI DART Lung Health Program (Innovate UK grant 40255), and the EPSRC Centre for Doctoral Training in Health Data Science (EP/S02428X/1).

## Acknowledgements

Views expressed are those of the authors and not necessarily those of the PathLAKE Consortium members, the NHS, the UKRI, the NIHR, Innovate UK or the Department of Health. JR is an adjunct professor of the Ludwig Oxford Branch.

## Author contributions

YH: Writing - original draft, Visualisation, Validation, Software, Methodology, Formal analysis, Data curation, Conceptualisation. GB: Software, Methodology, Formal analysis, Data curation. KG and MF: Validation, Investigation, Conceptualisation. ED: Validation, Investigation, Resources, Data curation, Conceptualisation. BL, TZ, and ZL: Writing - original draft, Conceptualisation. DW and CV: Conceptualisation. JR: Methodology, Project Management, Conceptualisation.

## Competing interests

The authors declare that they have no competing financial interests.

## Supplementary Information

The Supplementary Information provides additional cohort details for the ***FOCUS*** external validation set (Supplementary Table S1) and extended analyses that support the main results. Supplementary Figure S1 reports balanced accuracy (BACC) under different feature-compression levels. Supplementary Figure S2 quantifies spatial overlap of significant attention influx/efflux regions across compression settings and across PFMs, and Supplementary Figure S3 visualises cross-model conversion between influx and efflux assignments. Supplementary Figure S4 summarises class-wise distributions of attention influx/efflux clusters, Supplementary Figure S5 characterises slide-wise prevalence of influx phenotypes, and Supplementary Figure S6 provides task-agnostic cluster statistics and cross-model alignment results. Supplementary Figure S7 shows representative tile clusters with LLM-generated histomorphological descriptions, Supplementary Figure S8 links these clusters to their spatial context on tissue, and Supplementary Figure S9 presents representative local efflux-to-influx transitions with corresponding LLM narratives. Finally, Supplementary Figure S10, Supplementary Figure S11, and Supplementary Figure S12 report word-cloud and term-frequency analyses of medical LLM outputs for ***Ovarian-Bev-Resp, FOCUS-KRAS***, and ***Colitis-Marsh***, respectively.

## Footnotes

1 https://openslide.org/

2 https://pypi.org/project/opencv-python/

3 https://scikit-learn.org/stable/

4 https://chatgpt.com/

5 https://huggingface.co/microsoft/llava-med-v1.5-mistral-7b

6 https://amueller.github.io/word_cloud

7 https://huggingface.co/docs/transformers

8 https://matplotlib.org/

9 https://plotly.com/

10 https://seaborn.pydata.org/

11 https://github.com/konstantint/matplotlib-venn

## Notes

### Competing Interest Statement

The authors have declared no competing interest.

### Funding Statement

Authors were supported as: CV and JR were supported by the PathLAKE Centre of Excellence for digital pathology and artificial intelligence, which is funded by the Data to Early Diagnosis and Precision Medicine strand of the HM Government's Industrial Strategy Challenge Fund, managed and delivered by Innovate UK on behalf of UK Research and Innovation (UKRI) (Grant ref: 104689/application number 18181). YH and JR were supported by Novo Nordisk A/S - Big Data Institute (BDI), University of Oxford collaboration project. CV was supported by the NIHR Oxford Biomedical Research Centre. GB was supported by Fergus Gleeson's A2 research funds, UKRI DART Lung Health Program (Innovate UK grant 40255), and the EPSRC Centre for Doctoral Training in Health Data Science (EP/S02428X/1).

### Author Declarations

All the datasets used in your study had been de-identified prior to use in this study.

### Summary of Updates

All illustrations have been replaced with high-resolution (HD) versions.

