## Supplementary dataset information and results for "Harness Behavioural Analysis for Unpacking the Bio-Interpretability of Pathology Foundation Models"

### A Supplementary Information of FOCUS Cohort

The background of FOCUS trial: FOCUS was a large UK-based randomised controlled trial comparing different strategies of sequential or combination therapies of 5FUFA with or without oxaliplatin or irinotecan as first- or second-line therapies in patients with newly diagnosed advanced colorectal cancer.

The data acquisition protocol for FOCUS cohort: As part of the *S:CORT* program, patients with available formalin-fixed paraffin-embedded (FFPE) blocks of the primary CRC were selected from the FOCUS randomised clinical trial. Serial sections were cut from one representative block for H&E staining followed by four unstained sections for RNA extraction, a second H&E and eight unstained sections for DNA extraction. Glass H&E slides were rereviewed by an expert gastrointestinal pathologist and tumour tissue with the associated intratumoural stroma was annotated and used to guide RNA and DNA extractions from the first and second H&E respectively. No tumour microdissection was performed. Regions of extensive necrosis and non-tumour tissue were excluded according to standard practice for downstream molecular tumour profiling. RNA expression microarrays (Xcel array, Affymetrix) and DNA target capture (SureSelect, Agilent) followed by NGS sequencing (Illumina) were applied in this order. All H&E slides were scanned at high resolution on an Aperio scanner at a total magnification of 20x. Digital slides were re-reviewed by a second gastrointestinal pathologist and tumour annotations were traced to generate region annotations. Areas containing folds or debris were excluded by digital annotation. Clinical data was retrieved from the trial database and sidedness was extracted from pathological reports. Slides with technical failure of the staining or scanning procedure were excluded from further analysis. The population characteristics of FOCUS external validation set used in this paper are listed in Table S1.

**Table S1.** Population characteristics of FOCUS external validation set.

| **Characteristic** | **N = 100[1]** |
|---|---|
| **KRAS** | |
| Mut | 56 (56%) |
| Wt | 44 (44%) |
| **GENDER** | |
| Male | 63 (63%) |
| Female | 37 (37%) |
| **AGE** | 64 (57, 71) |
| **SIDEDNESS** | |
| Right | 39 (42%) |
| Left | 53 (58%) |
| Missing | 8 |
| **DISTANT METASTASIS** | |
| Synchronous | 64 (64%) |
| Metachronous | 36 (36%) |
| **TREATMENT ARM** | |
| MdG → Ir | 44 (44%) |
| MdG → IrMdG | 23 (23%) |
| MdG → OxMdG | 15 (15%) |
| OxMdG | 18 (18%) |

[1] n (%); Median (IQR)

### B Supplementary Figures

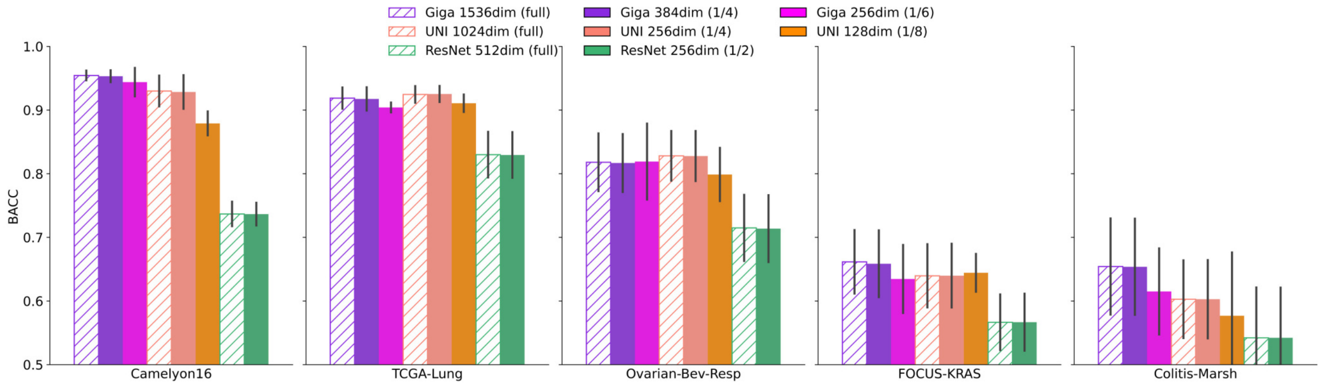


**Figure S1.** BACC performance of the foundation model features with different compression levels across downstream tasks. Diagonal hatching denotes uncompressed (original) features; solid colours indicate varying degrees of compression.

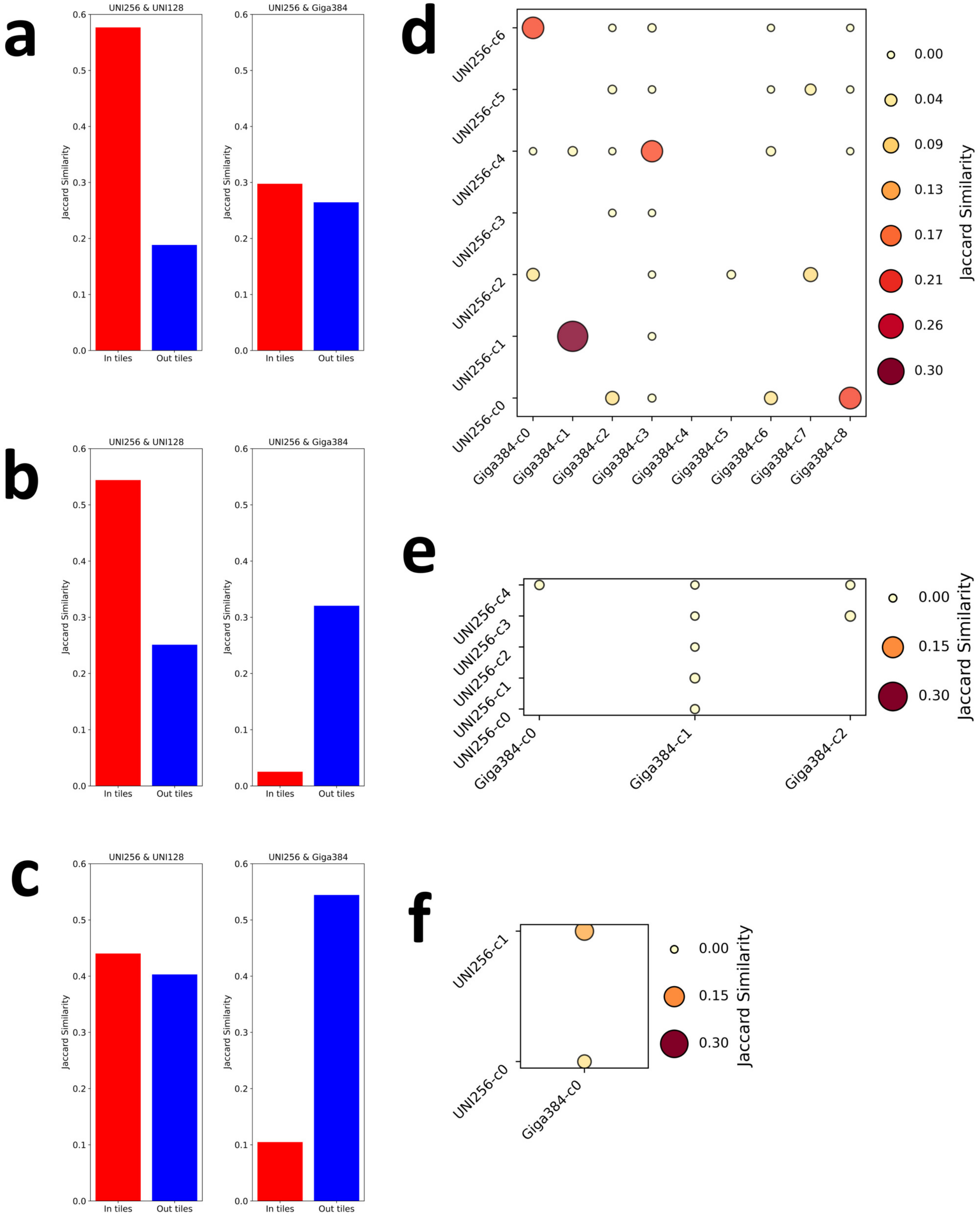


**Figure S2. Spatial overlap of attention influx phenotypes based on different PFMs. a - c**, Jaccard similarity of significant attention influx and efflux regions across feature sets derived from different compression levels of the same foundation model (UNI) and across different foundation models (**a**: Ovarian-Bev-Resp; **b**: FOCUS-KRAS; **c**: Colitis-Marsh). **d - f**, Overlap between attention influx phenotype clusters derived from UNI and prov-GigaPath features, assessed via pairwise similarity (**d**: Ovarian-Bev-Resp; **e**: FOCUS-KRAS; **f**: Colitis-Marsh).

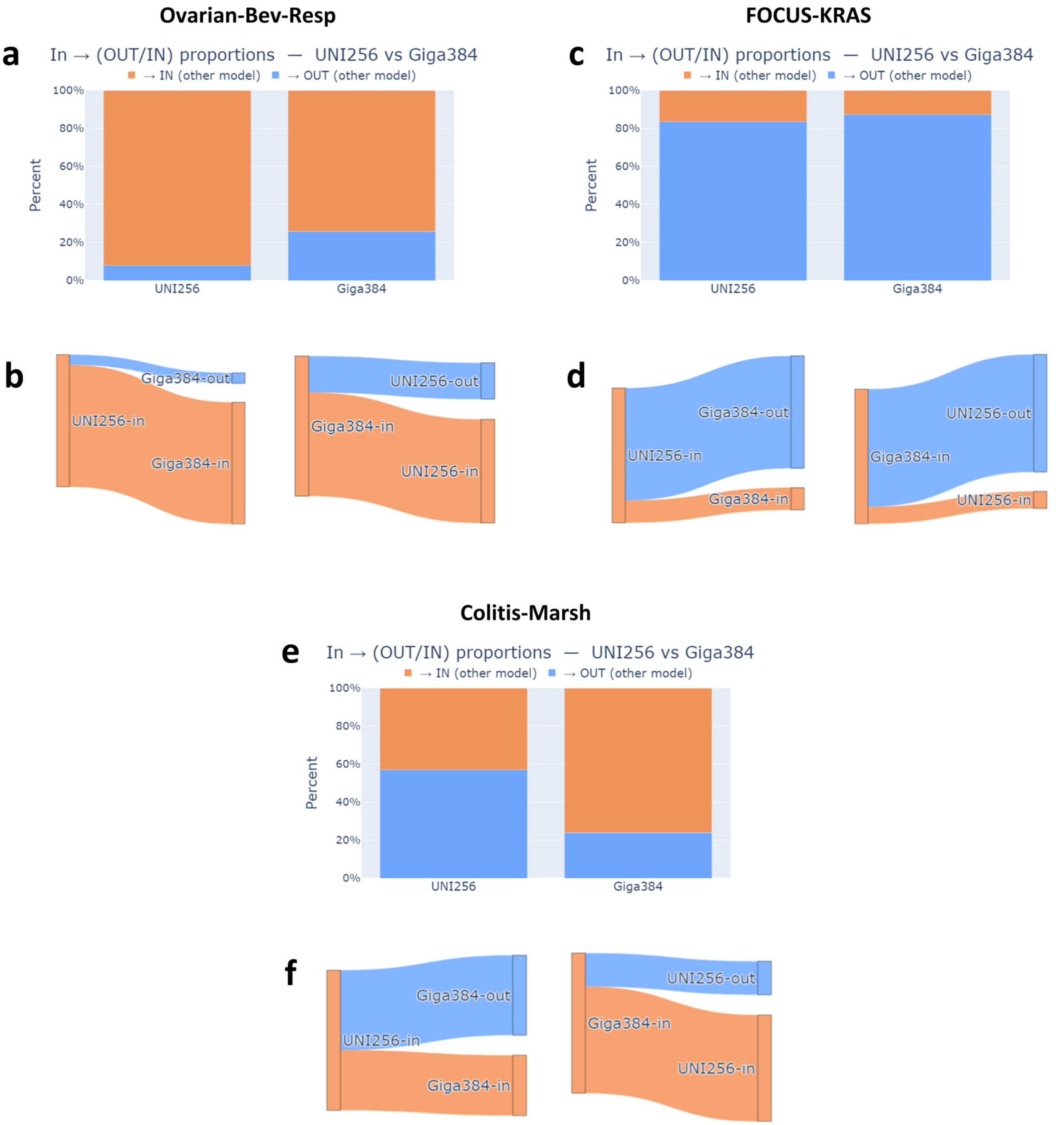


**Figure S3.** **Cross-model conversion of attention influx and efflux across datasets. a,c,e,** Butterfly plots showing the proportion of tiles marked as significant attention influx by one PFM (UNI-256 or GigaPath-384) that either remain influx or convert into efflux regions when interpreted by the other PFM. A higher conversion rate indicates stronger divergence, or even opposite interpretative behaviour, between the two models. **b,d,f,** Corresponding Sankey diagrams illustrating the same conversions, with flows connecting influx (orange) and efflux (blue) assignments across PFMs. Results are shown for three representative downstream tasks: (**a,b**) Ovarian-Bev-Resp, (**c,d**) FOCUS-KRAS, and (**e,f**) Colitis-Marsh.

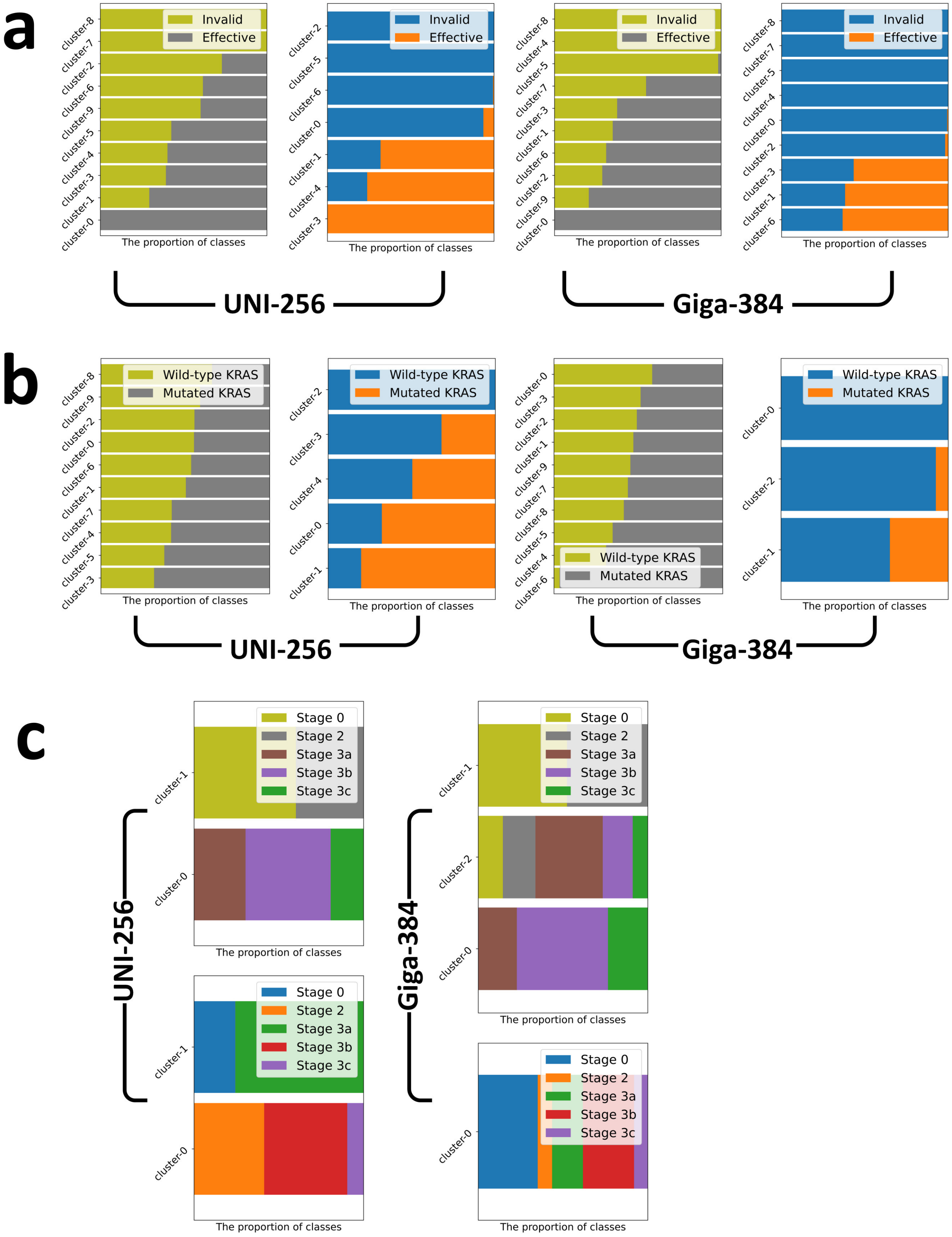


**Figure S4. Distribution of attention influx and efflux clusters across different classes. a,** In the task of Ovarian-Bev-Resp. For different PFMs, left: clusters of attention efflux, right: clusters of attention influx. **b,** In the task of OCUS-KRAS. Left: clusters of attention efflux, right: clusters of attention influx. **c,** In the task of Colitis-Marsh. For different PFMs, top: clusters of attention efflux, bottom: clusters of attention influx.

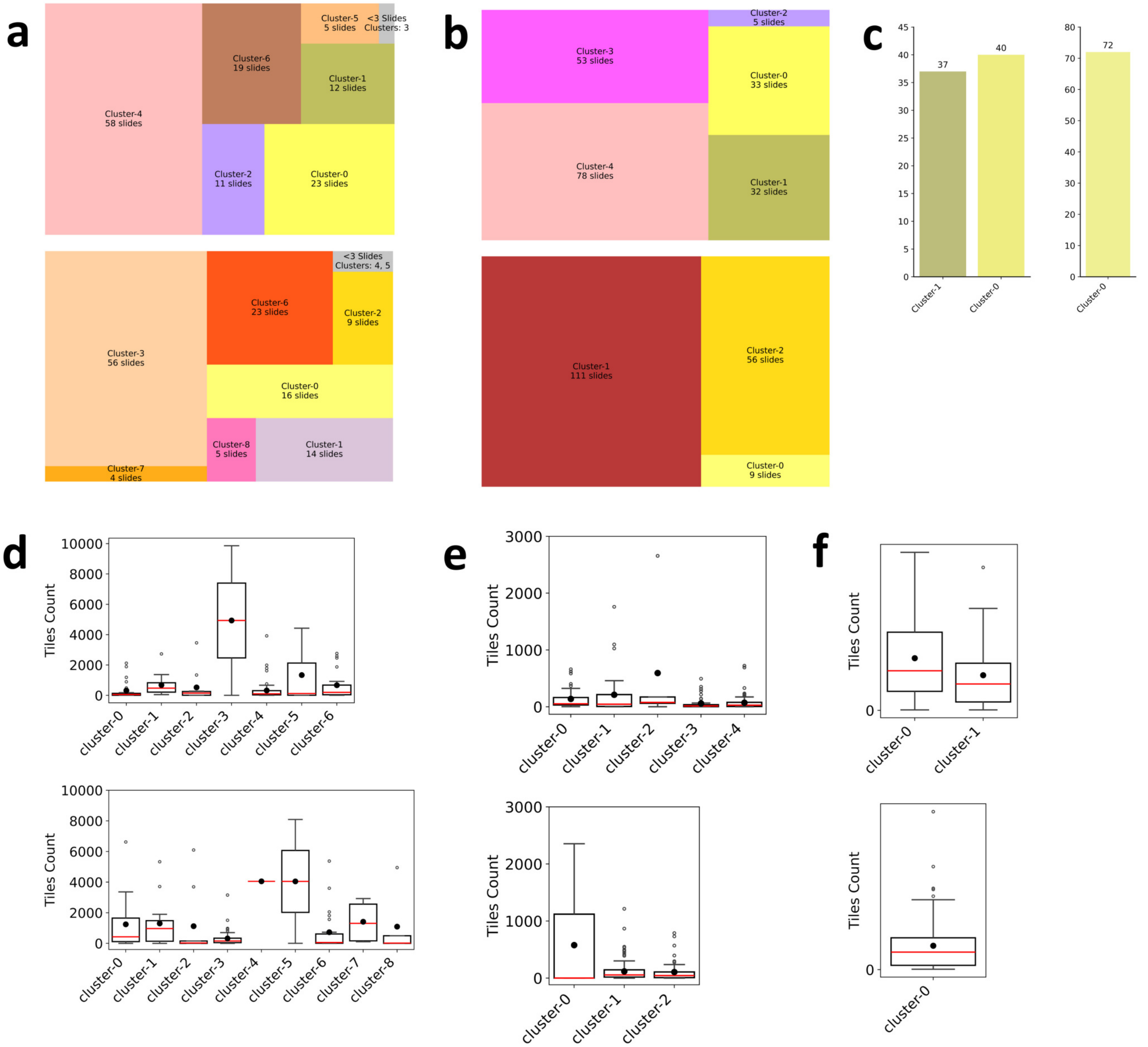


**Figure S5. Task-specific distribution of attention influx phenotypes. a** and **b**, Treemaps showing the number of slides in which each attention influx phenotype cluster appears, for both UNI (top) and Giga features (bottom) (**a**: Ovarian-Bev-Resp; **b**: FOCUS-KRAS). **c**, Bar chart to show the number of slides containing each attention influx phenotype cluster, for both UNI (left) and Giga features (right), on task - Colitis-Marsh. **d** - **f**, Distribution of the number of attention influx phenotype clusters per slide, on different tasks (**d**: Ovarian-Bev-Resp; **e**: FOCUS-KRAS; **f**: Colitis-Marsh).

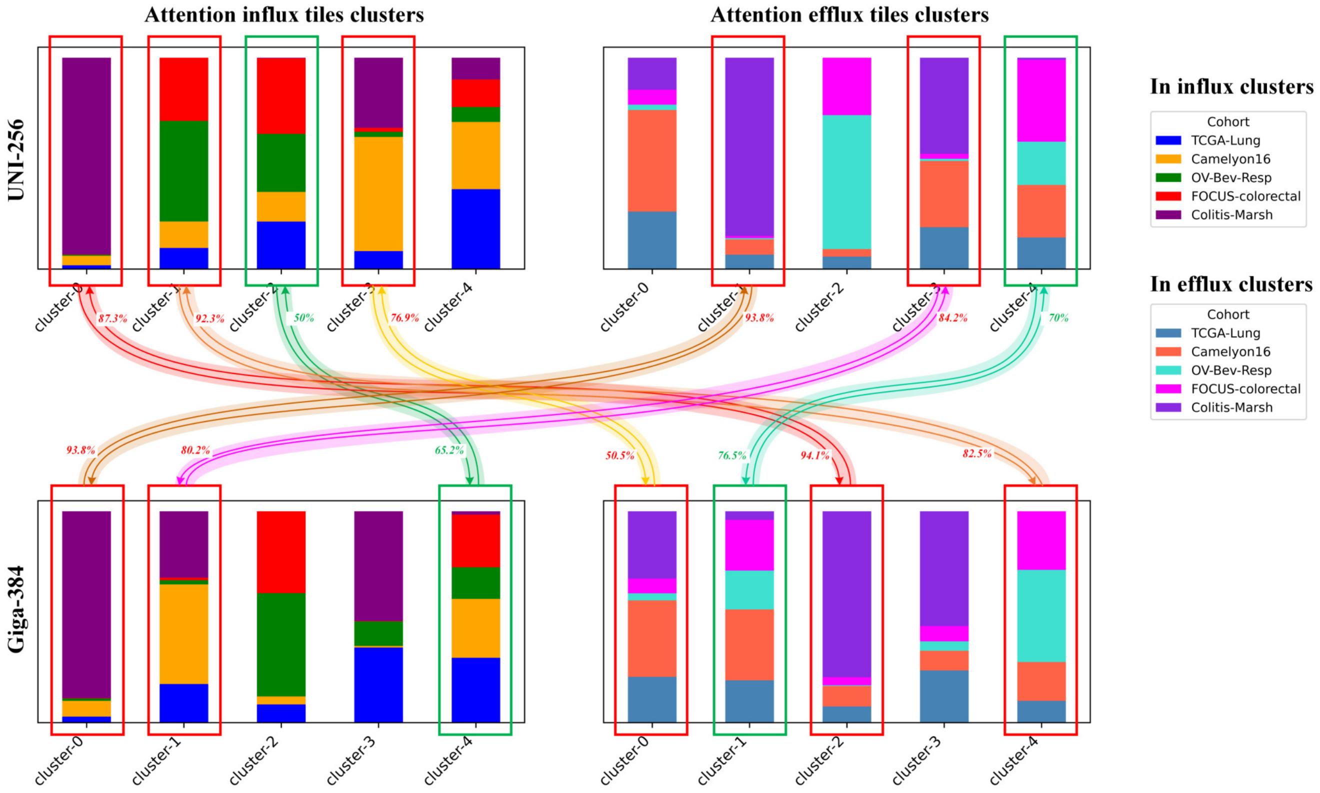


**Figure S6. Cluster distribution statistics of task-agnostic significant attention influx/efflux features.** Proportional distribution of attention-related phenotype clusters across datasets, aggregated over multiple downstream tasks. Highlighted cluster pairs passed bidirectional spatial alignment thresholds (fit > 50%). Warm-coloured curves indicate mismatched attention direction between UNI and Giga features (e.g. influx vs. efflux), while cool-coloured curves indicate consistent attention behaviour.

Initial prompt

You are a pathologist and need to review some image analysis results. I will continue to provide you with a set of 256 x 256 pathology images. These are H&E-stained patched biopsies of ovarian cancer patients. You will be asked to help review each set of images and describe the histological phenotypes they exhibit.

Q&A about sample tiles for Influx cluster-0 (c-0)

**UNI**

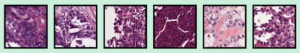

Could you tell me what histological phenotypic features are presented in these pathological patches as a whole?

These pathological image patches collectively show features consistent with high-grade epithelial ovarian carcinoma, most likely high-grade serous carcinoma. The main histological phenotypes include:
Nuclear Atypia: Enlarged, pleomorphic nuclei with irregular contours and prominent nucleoli are evident across several patches, reflecting high-grade cytological atypia.
High Mitotic Activity: Frequent mitotic figures are present, indicating rapid proliferative activity of the tumour cells.
Loss of Normal Architecture: The images demonstrate solid nests and sheets of tumour cells with poorly defined glandular structures, replacing the normal ovarian stroma.
Hyperchromasia: The nuclei are densely stained (hyperchromatic), further supporting malignant transformation.
Stromal Desmoplasia and Necrosis: Some patches suggest stromal reaction and areas of necrotic debris, characteristic of aggressive tumour behaviour.
Cellular Cohesion and Stratification: Tumour cells are arranged in disorganised layers with loss of polarity, consistent with invasive carcinoma.
Overall, these features represent poorly differentiated, aggressive tumour morphology, compatible with high-grade serous ovarian carcinoma, which is the most common subtype encountered in clinical pathology.

**Giga**

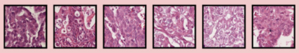

Could you tell me what histological phenotypic features are presented in these pathological patches as a whole?

These pathological patches collectively demonstrate a high-grade carcinoma phenotype with marked pleomorphism and solid growth patterns. The key histological features include:
Marked Nuclear Atypia: Enlarged, irregular, and hyperchromatic nuclei with conspicuous nucleoli. There is significant variation in nuclear size and shape, a hallmark of high-grade malignancy.
Loss of Cohesive Architecture: The tumour cells form irregular, poorly defined nests and sheets, with areas of discohesion and scattered single cells, indicating invasive behaviour.
Prominent Mitotic Activity: Several fields suggest brisk mitotic activity, including atypical mitoses, reflecting rapid proliferation.
Epithelial Clustering with Necrosis: Tumour nests are intermixed with eosinophilic necrotic debris and apoptotic bodies, consistent with high cellular turnover.
Cytoplasmic Variation: The tumour cells show moderate, sometimes vacuolated or eosinophilic cytoplasm, contributing to morphological heterogeneity.
Possible Papillary/Glandular Differentiation: In some regions, there are rudimentary gland-like lumina, though largely obscured by solid growth and pleomorphism.
Overall, these features point towards a high-grade serous ovarian carcinoma with solid/discohesive growth, marked nuclear pleomorphism, and high mitotic activity, highlighting its aggressive biological behaviour.

Q&A about sample tiles for influx cluster-1 (c-1)

**UNI**

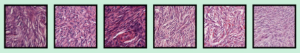

Could you tell me what histological phenotypic features are presented in these pathological patches as a whole?

These pathological patches overall display features of spindle cell morphology. The main histological phenotypes observed include:
Spindle-Shaped Cells, which are the majority of cells, are elongated with tapered ends, arranged in fascicles or interlacing bundles, characteristic of stromal or mesenchymal differentiation.
Storiform and Fascicular Patterns: Cells are aligned in sweeping fascicles and sometimes whorled or storiform arrangements, typical of stromal tumours.
Mild to Moderate Nuclear Atypia: Nuclei are elongated, hyperchromatic, but less pleomorphic compared to high-grade carcinoma. Some nuclei appear blunt-ended or cigar-shaped.
Collagenous/Fibrous Stroma: There is abundant eosinophilic extracellular matrix, consistent with fibrous stroma deposition.
Low Mitotic Activity: Compared to the previous malignant epithelial set, mitotic figures appear less frequent, suggesting a relatively lower proliferative index.
Absence of Glandular/Epithelial Structures: Unlike carcinoma, these patches lack gland formation, papillary structures, or cohesive epithelial nests, reinforcing a mesenchymal phenotype.
Taken together, these features are most compatible with ovarian stromal or sex cord–stromal tumour morphology (such as fibroma, thecoma, or related spindle cell lesions). They reflect a mesenchymal phenotype rather than epithelial carcinoma.

**Giga**

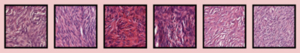

Could you tell me what histological phenotypic features are presented in these pathological patches as a whole?

These pathological patches overall demonstrate a stromal/spindle-cell phenotype with characteristic features of fibrous or mesenchymal tissue. The main histological findings include:
Spindle-Shaped Cells: The majority of cells are elongated with tapered ends, arranged in bundles and fascicles, typical of stromal or mesenchymal morphology.
Interlacing Fascicular Architecture: The cells form sweeping, interwoven fascicles and whorled patterns, a hallmark of fibrous stroma or sex cord–stromal tumour morphology.
Dense Collagen Deposition: Abundant eosinophilic extracellular matrix, likely collagen, is present, indicating a fibrotic component.
Relatively Low Nuclear Atypia: Compared with high-grade carcinoma patches, nuclei here are less pleomorphic, though elongated and hyperchromatic. This suggests a stromal rather than epithelial malignant phenotype.
Scant Cytoplasm: The cells have elongated nuclei with little surrounding cytoplasm, reinforcing a fibroblastic/myofibroblastic appearance.
Lack of Epithelial Structures: No glandular, papillary, or solid epithelial nests are evident, separating this morphology from carcinoma and pointing towards a stromal lesion.
Taken together, these features represent a stromal/mesenchymal phenotype, such as ovarian fibroma, thecoma, or a fibrosarcoma-like morphology in more atypical contexts.

Q&A about sample tiles for cluster-2

**UNI**

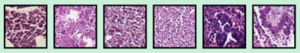

Could you tell me what histological phenotypic features are presented in these pathological patches as a whole?

These pathological patches overall exhibit inflammatory and tumour-associated microenvironmental features with a high density of small, round cells mixed with malignant epithelial elements. The key histological phenotypes include:
Prominent Lymphoid Infiltration: Numerous small round lymphocyte-like cells with dark nuclei and scant cytoplasm are present, suggesting a dense immune response within the tumour microenvironment.
Tumour Cells with Pleomorphism: Amidst the inflammatory infiltrates, clusters of larger malignant cells with hyperchromatic, irregular nuclei and moderate cytoplasm are visible, indicating carcinoma elements.
Discohesive Growth Pattern: Some areas show scattered, loosely arranged malignant cells rather than cohesive nests or glands, reflecting an infiltrative pattern.
Necrotic and Apoptotic Debris: Several fields reveal areas of cell death, with eosinophilic debris and nuclear fragmentation, consistent with tumour necrosis and apoptosis.
Reactive Stromal Changes: The surrounding stroma demonstrates oedematous changes and vascular congestion, possibly linked to tumour-induced inflammatory reaction.
Mixed Cellular Population: Besides malignant epithelial cells, there are abundant immune cells, occasional plasma cells, and possibly histiocytes, contributing to a heterogeneous tumour-immune microenvironment.
Taken together, these features represent a carcinoma with intense lymphoid infiltration and tumour necrosis, which is commonly seen in high-grade ovarian carcinomas with strong host immune responses.

Q&A about sample tiles for cluster-3

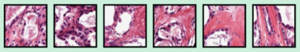

Could you tell me what histological phenotypic features are presented in these pathological patches as a whole?

These patches as a whole show desmoplastic stromal reaction with embedded malignant epithelial cells, a phenotype often seen in invasive ovarian carcinoma. The key histological features include:
Prominent Fibrous/Collagenous Stroma: Dense pink eosinophilic bundles of collagen are present throughout the images, consistent with desmoplasia, which is a stromal response to invasive carcinoma.
Scattered Malignant Epithelial Cells: Within and around the fibrous tissue, tumour cells with hyperchromatic, pleomorphic nuclei can be seen. These cells often appear irregular and embedded within the fibrous matrix.
Stromal-Tumour Interaction: The tumour cells appear to infiltrate and entrap the stroma, suggesting invasive growth rather than expansion in well-formed nests or glands.
Nuclear Atypia: The epithelial cells show irregular nuclear contours, variation in size and shape, and prominent nucleoli, features consistent with high-grade carcinoma.
Fibroblastic/Reactive Stromal Cells: In addition to collagen deposition, elongated fibroblastic cells are intermixed, further supporting a desmoplastic reaction.
Occasional Necrotic/Apoptotic Debris: Some areas suggest degenerative changes and possible necrosis, reflecting tumour aggressiveness.
Taken together, these images demonstrate a desmoplastic carcinoma phenotype, characterised by invasive malignant epithelial cells inducing a robust fibrous stromal response. This pattern is particularly common in high-grade serous ovarian carcinoma and signifies invasive disease.

**Giga**

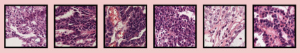

Could you tell me what histological phenotypic features are presented in these pathological patches as a whole?

These pathological patches as a whole demonstrate features of high-grade epithelial ovarian carcinoma with marked heterogeneity. The histological phenotypes observed include:
Solid Sheets and Nests of Tumour Cells: Most images show densely packed tumour cells with limited glandular or papillary formation, consistent with poorly differentiated carcinoma.
Nuclear Atypia: Enlarged, irregular, hyperchromatic nuclei with conspicuous nucleoli. There is marked pleomorphism and variability in nuclear size and shape.
High Mitotic Activity: Evidence of brisk proliferation with numerous mitotic figures, suggestive of aggressive tumour behaviour.
Apoptosis and Necrosis: Some areas show apoptotic bodies and cytoplasmic degeneration, indicative of high cellular turnover.
Stromal Desmoplasia: Certain patches show fibrous stroma with infiltrating malignant cells, reflecting invasive growth.
Architectural Diversity: While most areas are solid, some exhibit focal gland-like structures or cribriform patterns, highlighting intratumoural heterogeneity.
Overall, these features are most consistent with high-grade serous ovarian carcinoma, a subtype defined by pleomorphic nuclei, high mitotic index, apoptosis, and invasive architecture.

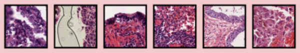

Could you tell me what histological phenotypic features are presented in these pathological patches as a whole?

These patches as a whole demonstrate a heterogeneous spectrum of histological phenotypes:
Epithelial proliferations: Several patches show cohesive epithelial nests and sheets with enlarged, hyperchromatic nuclei, consistent with malignant epithelial components of ovarian carcinoma.
Papillary and glandular structures: Some areas show columnar epithelial lining with stratified nuclei, suggesting papillary/glandular differentiation, a frequent feature of serous ovarian carcinoma.
Inflammatory infiltration: There are foci with dense lymphocytic and mixed inflammatory cell infiltration, indicating host immune response within the tumour microenvironment.
Necrosis and degenerative changes: Certain regions contain necrotic debris and apoptotic bodies, typical of high-grade carcinomas with rapid turnover.
Stromal reaction: Spindle-shaped fibroblasts and desmoplastic stroma are evident in some fields, reflecting stromal-epithelial interaction.
Haemorrhage and vascular congestion. Patches reveal erythrocyte extravasation and congested vessels, indicating haemorrhagic changes often seen in high-grade ovarian cancer.
Overall phenotype:
These images are consistent with high-grade serous carcinoma morphology, showing malignant epithelial nests with nuclear atypia, papillary/glandular differentiation, stromal desmoplasia, inflammatory response, and necrotic/haemorrhagic areas, which is a composite picture of an aggressive ovarian carcinoma microenvironment.

Q&A about sample tiles for cluster-4

**UNI**

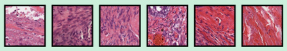

Could you tell me what histological phenotypic features are presented in these pathological patches as a whole?

These pathological patches overall display a stromal-dominant phenotype with desmoplasia and scattered malignant elements, suggestive of an invasive carcinoma with strong stromal reaction. The main histological features include:
Dense Collagenous Stroma: Several patches show abundant eosinophilic collagen bundles (intensely pink/red staining), arranged in parallel or haphazard fashion, characteristic of desmoplastic reaction.
Spindle-Shaped Stromal Cells: Within the fibrotic stroma, elongated fibroblastic or myofibroblastic cells are evident, consistent with stromal activation.
Scattered Malignant Epithelial Cells: Some areas (e.g., third patch) show clusters of atypical epithelial cells with pleomorphic, hyperchromatic nuclei, embedded within the stromal background.
Vascular and Haemorrhagic Elements: A few regions reveal congested vessels and red blood cell extravasation, indicating tumour-related angiogenesis and stromal remodelling.
Inflammatory Cell Infiltration: Occasional lymphocytes and histiocytes are interspersed within the stroma, reflecting immune–tumour interaction.
Heterogeneous Tissue Composition: The variation between images (from collagen-rich fibrous tissue to mixed tumour–stromal zones) highlights the complexity of the tumour microenvironment.
Taken together, these features represent a desmoplastic ovarian carcinoma microenvironment, where the tumour induces extensive fibrotic stromal response, often associated with invasiveness and aggressive tumour biology.

Q&A about sample tiles for cluster-5

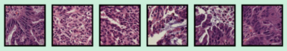

Could you tell me what histological phenotypic features are presented in these pathological patches as a whole?

These pathological patches as a whole show features of high-grade epithelial carcinoma with marked nuclear atypia and architectural disorganisation, typical of aggressive ovarian carcinoma. The main histological phenotypes include:
Solid Sheets and Nests of Tumour Cells: The tumour cells are arranged in dense, cohesive clusters with little evidence of gland formation, reflecting a poorly differentiated growth pattern.
Marked Nuclear Pleomorphism: Nuclei are enlarged, irregular, and hyperchromatic, with prominent nucleoli and variation in size and shape, consistent with high-grade cytological atypia.
Loss of Polarity and Stratification: Cells are disorganised, with loss of normal epithelial orientation, contributing to a chaotic architecture.
High Mitotic Activity: Numerous mitotic figures and atypical mitoses can be appreciated, indicative of rapid proliferation.
Apoptosis and Necrosis: Some fields show apoptotic bodies and degenerative changes, reflecting high cellular turnover.
Infiltrative Pattern: At the periphery, tumour cells appear to invade surrounding stroma, further confirming their malignant behaviour.
Overall, these features are characteristic of high-grade serous ovarian carcinoma, the most common and aggressive subtype, notable for its poorly differentiated morphology, high proliferative index, and extensive nuclear atypia.

Q&A about sample tiles for cluster-5

**Giga**

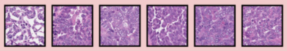

Could you tell me what histological phenotypic features are presented in these pathological patches as a whole?

Across these patches, the histological phenotypic features are characterised by:
High nuclear density: The fields are tightly packed with cells, leaving little extracellular space.
Nuclear atypia and pleomorphism: Many nuclei vary in size and shape, some enlarged and hyperchromatic.
Prominent nucleoli: In several cells, nucleoli are visible, reflecting increased transcriptional activity.
Cytoplasmic clearing and variation: Some cells exhibit distinct cytoplasmic borders with irregular clearing, while others have more granular cytoplasm.
Mitotic activity: The nuclear morphology suggests increased proliferative activity, with scattered mitotic figures.
Loss of normal tissue organisation: Unlike normal glandular or stromal architecture, these patches show a disordered, sheet-like or solid growth pattern.
Possible necrotic/apoptotic areas: Some pale regions with fragmented nuclear material may indicate focal necrosis or apoptosis.
Taken together, these features suggest a high-grade malignant phenotype with aggressive behaviour, consistent with poorly differentiated carcinoma or other high-grade neoplasms, depending on the tissue of origin.

Q&A about sample tiles for cluster-6

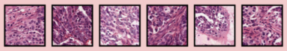

Could you tell me what histological phenotypic features are presented in these pathological patches as a whole?

These pathological patches as a whole present the following histological phenotypic features:
Cellular atypia: Cells display enlarged, irregular, and hyperchromatic nuclei with prominent nucleoli, consistent with neoplastic transformation.
Increased nuclear-to-cytoplasmic ratio: Many cells show dense nuclear material relative to scant cytoplasm, a hallmark of malignancy.
Pleomorphism: There is noticeable variability in nuclear size and shape across the fields.
Mitotic activity: Frequent mitotic figures are visible, indicating a high proliferative index.
Loss of normal tissue architecture: The organisation of epithelial structures appears disrupted, replaced by disordered clusters of atypical cells.
Stromal reaction: In some areas, desmoplastic stroma and fibrous tissue infiltration are present, reflecting tumour–stroma interaction.
Possible invasion: Clusters of atypical cells appear to infiltrate the surrounding stroma, suggesting invasive growth.
Taken together, these features are characteristic of a high-grade malignant epithelial tumour, such as carcinoma, with evidence of aggressive behaviour (pleomorphism, mitotic activity, invasion).

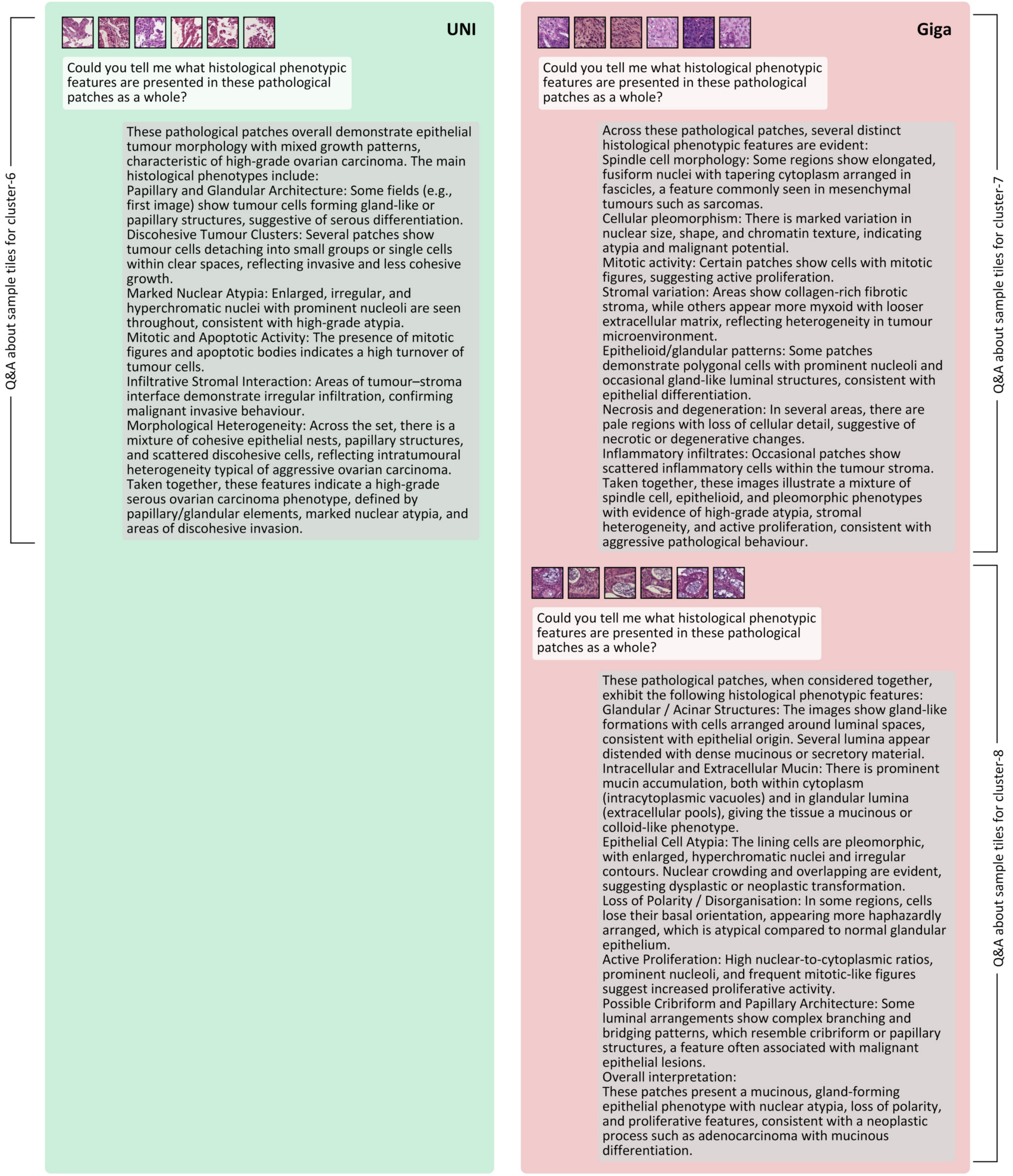


**Figure S7. LLM-based histopathological descriptions of attention influx clusters (Ovarian-Bev-Resp).** Representative examples of tile clusters with significant attention influx identified from UNI and prov-GigaPath embeddings in the Ovarian-Bev-Resp dataset. Each panel shows the initial prompt followed by LLM-generated morphological descriptions: the left column corresponds to representative clusters from UNI, and the right column corresponds to those from prov-GigaPath. These examples illustrate how a locally deployed LLM can characterise the distinct histological phenotypes emphasised by different foundation models. The figure is divided across four pages but shares a single caption.

Initial prompt

You are a pathologist, and you need to help me review some pathology image analysis results. These images are H&E-stained pathology images from patients with ovarian cancer. I will now show you images of sections of a large tissue, some of which are marked with coloured grids. You need to focus on the areas marked with specific colours and describe the morphological phenotypes they reveal.

Q&A about sample tissue area for cluster-0

**UNI**

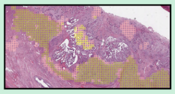

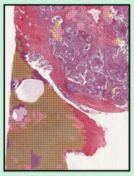

Tell me what morphological phenotype the exactly Yellow grid indicated area in the tissues given to you shows?

The yellow grid-marked regions in both ovarian cancer H&E slides highlight areas with the following key morphological phenotypes:
Tumour epithelial proliferation: The yellow-marked areas correspond to glandular or papillary epithelial structures, showing malignant epithelial proliferation. These tumour cells form complex gland-like spaces with irregular lumina. Nuclei appear enlarged, hyperchromatic, and crowded, with evidence of stratification. This is consistent with high-grade serous carcinoma morphology, which commonly shows papillary, cribriform, or solid growth patterns.
Desmoplastic stromal reaction: Surrounding the epithelial nests, there is fibrous stroma with increased cellularity, indicating stromal desmoplasia, a typical host response to invasive carcinoma. The yellow grids tend to highlight the interface between tumour epithelium and reactive stroma, emphasising tumour–stroma interaction zones.
Features of invasion: The epithelial clusters within yellow-marked areas appear irregular and infiltrative, extending into the fibrotic stroma, supporting the diagnosis of invasive carcinoma rather than a borderline tumour.
In summary:
The yellow grid regions capture areas of invasive malignant epithelial proliferation with papillary/glandular morphology, nuclear atypia, and desmoplastic stromal reaction, characteristic of high-grade serous carcinoma of the ovary.

**Giga**

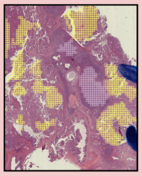

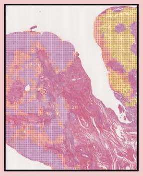

Tell me what morphological phenotype the exactly Bright Yellow grid indicated area in the tissues given to you shows?

The bright yellow grid–marked regions in these ovarian cancer H&E slides correspond to a morphological phenotype defined by:
Dense malignant epithelial proliferation: These areas are composed of closely packed tumour epithelial cells with reduced intervening stroma. The architecture shows solid nests, papillary fronds, and irregular glandular structures, typical of high-grade serous carcinoma.
Pronounced nuclear atypia: Cells display enlarged, pleomorphic, hyperchromatic nuclei with prominent nucleoli. Nuclear crowding and overlapping are evident, with loss of polarity.
High mitotic activity: Frequent mitotic figures are present, sometimes atypical, consistent with an aggressive proliferative compartment.
Evidence of invasion: The bright yellow regions infiltrate into adjacent stroma, disrupting normal tissue boundaries. Some tumour nests show irregular contours with surrounding desmoplastic reaction.
In summary, the bright yellow grid areas represent viable, highly proliferative tumour epithelium with papillary/solid/glandular architecture, marked nuclear atypia, and stromal invasion, characteristic of the active invasive component of high-grade serous carcinoma.

Q&A about sample tissue area for cluster-1

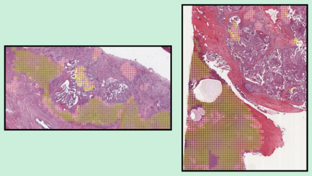


Tell me what morphological phenotype the exactly Olive Green grid indicated area in the tissues given to you shows?

The olive green grid-marked regions in these ovarian cancer H&E slides show a different histological phenotype compared to the yellow-marked tumour foci. Specifically:
Predominantly stromal/fibrous tissue:
The olive green areas correspond mainly to fibrous stroma with abundant extracellular matrix and elongated spindle-shaped fibroblasts. The stroma appears dense and collagenised, showing a desmoplastic reaction associated with the invasive carcinoma. In several zones, the stroma is relatively acellular compared to the adjacent tumour epithelium, emphasising its supportive rather than proliferative nature.
Reduced epithelial component:
Unlike the yellow regions, the olive green grids do not highlight dense tumour cell nests, but instead areas with sparse or absent epithelial structures. Occasional small tumour glands or scattered malignant cells may be embedded within the stroma, but the overall pattern is stromal-dominant.
Possible inflammatory and vascular elements:
Some olive green–highlighted areas contain inflammatory infiltrates (lymphocytes and plasma cells) interspersed within the fibrous tissue. Small blood vessels can also be identified, reflecting stromal vascularisation associated with tumour invasion.
In summary:
The olive green grid regions primarily represent fibrotic/desmoplastic stroma with variable inflammatory infiltrates and vascular elements, reflecting the tumour microenvironment rather than direct epithelial tumour proliferation. This contrasts with the yellow-marked areas, which show malignant epithelial proliferation.

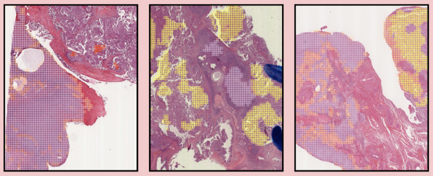


Tell me what morphological phenotype the exactly Light Purple grid indicated area in the tissues given to you shows?

The light purple grid–marked regions in these ovarian cancer H&E slides show a morphological phenotype characterised by:
Predominantly fibrous stroma:
These areas are composed largely of dense collagenous connective tissue with relatively few epithelial tumour cells. The stroma appears pale, eosinophilic, and homogenised, typical of desmoplastic or fibrotic tissue reaction.
Low tumour cellularity:
Compared to the adjacent bright yellow tumour-rich zones, the light purple areas contain sparse or scattered malignant cells. Tumour nests may be small, fragmented, or even absent, leaving the field dominated by stromal elements.
Vascular and inflammatory components:
Within the fibrous stroma, there are small-calibre blood vessels and occasional lymphocytic infiltrates, consistent with tumour-associated stromal response.
Transitional zones: These regions often lie at the interface between highly cellular carcinoma foci and surrounding supportive tissue, representing areas where tumour and stroma interact most strongly.
In summary, the light purple grid areas represent stroma-dominant regions with dense fibrosis, sparse epithelial tumour cells, and occasional inflammatory/vascular elements, reflecting a desmoplastic stromal reaction associated with invasive carcinoma.

Q&A about sample tissue area for cluster-2

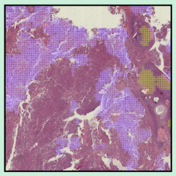

Tell me what morphological phenotype the exactly Purple grid indicated area in the tissues given to you shows?

The purple grid-marked areas in this ovarian cancer H&E section show:
Solid nests and sheets of tumour cells with little to no glandular or papillary lumen formation.
High nuclear atypia: enlarged, pleomorphic, hyperchromatic nuclei, often with prominent nucleoli.
Dense cellularity, with tumour cells packed closely together.
Frequent mitotic figures, indicating high proliferative activity.
Focal necrosis or degeneration may also be present within some purple-marked areas.
These features indicate a solid growth pattern of high-grade serous carcinoma, reflecting an aggressive malignant phenotype characterised by compact epithelial proliferation and marked cytological atypia.

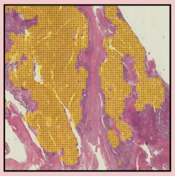

Tell me what morphological phenotype the exactly Bright Gold grid indicated area in the tissues given to you shows?

The bright gold grid–marked regions in this ovarian cancer H&E slide represent a morphological phenotype with the following features:
Fibrous/desmoplastic stroma predominance: These areas are mainly composed of dense eosinophilic collagenous tissue, with elongated fibroblasts. The stroma appears broad, band-like, and acellular in large portions, reflecting extensive fibrosis.
Low epithelial tumour component:
Unlike the yellow or bright yellow regions rich in carcinoma cells, the bright gold zones show very few or absent malignant epithelial nests. Tumour cells may be sparse and scattered, embedded within the fibrotic background.
Oedematous and hyalinised texture: Some areas demonstrate oedematous stroma with hyalinisation, giving a pale, homogenised appearance. This suggests a chronic stromal reaction rather than active epithelial proliferation.
Vascular elements:
Occasional thin-walled blood vessels traverse the fibrotic stroma, sometimes congested. No significant necrosis is evident within these zones.
In general, the bright gold grid areas correspond to fibrosis-dominant stromal regions with extensive collagen deposition, minimal tumour cell content, and scattered vascular elements, representing a strong desmoplastic stromal reaction rather than an epithelial tumour-rich compartment.

Q&A about sample tissue area for cluster-3

**UNI**

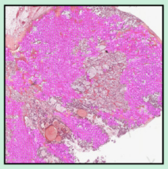

Tell me what morphological phenotype the exactly **Pink** grid indicated area in the tissues given to you shows?

The pink grid-marked regions in this ovarian cancer H&E section show the following morphological phenotypes characterised by:
Necrosis and degeneration: The pink-marked areas largely correspond to pale, eosinophilic, structureless zones with ghost outlines of tumour cells. These features are consistent with tumour necrosis, where viable tumour tissue has undergone cell death. Cellular details are lost, and only residual debris and karyorrhectic nuclear fragments may be seen.
Cystic/degenerative spaces: Within these zones, there are large empty or eosinophilic fluid-filled spaces and clefts, reflecting cystic degeneration. The surrounding tumour tissue shows abrupt transitions between viable carcinoma and necrotic zones.
Haemorrhage and vascular congestion: Some pink-marked regions contain red blood cell extravasation and congested vessels, suggesting associated haemorrhage within necrotic tissue.
In summary:
The pink grid areas represent extensive tumour necrosis with cystic degeneration and focal haemorrhage, reflecting a non-viable tissue phenotype often seen in aggressive, high-grade carcinomas due to rapid tumour growth outpacing vascular supply.

Q&A about sample tissue area for cluster-4

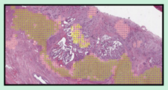

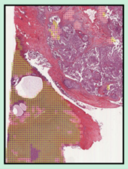

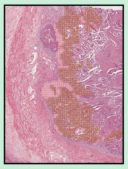

Tell me what morphological phenotype the exactly **Light Pink** grid indicated area in the tissues given to you shows?

The light pink grid-marked regions in these ovarian cancer H&E slides highlight areas with the following morphological phenotype:
Loose fibrotic stroma:
These regions predominantly show pale eosinophilic connective tissue with fewer tumour cells compared to adjacent carcinoma-rich areas. The stroma is less densely collagenised than the olive green-marked desmoplastic stroma, often appearing looser and more oedematous.
Scattered inflammatory infiltrates: Occasional lymphocytes and plasma cells are present within this stroma, suggesting an immune response. However, the density of inflammatory cells is relatively modest compared to reactive hotspots.
Transition zones: The light pink grid areas often lie at the interface between dense tumour nests and surrounding fibrotic tissue, functioning as “border” regions. In some places, small detached tumour clusters or single malignant cells are present within this looser stroma, reflecting early invasion.
So, the light pink grid areas represent stromal transition zones with loose fibrotic tissue, modest inflammatory infiltration, and occasional infiltrating tumour cells. They act as intermediate compartments between dense carcinoma and fully desmoplastic stroma.

**Giga**

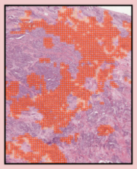

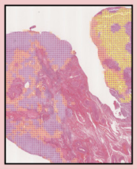

Tell me what morphological phenotype the exactly **Light Orange** grid indicated area in the tissues given to you shows?

The light orange grid–marked regions in these ovarian cancer H&E slides represent a morphological phenotype with the following features:
Loose fibrotic stroma with patchy tumour infiltration: These zones are dominated by pale eosinophilic stromal tissue that is less dense than the brown or bright gold stromal areas. There are scattered malignant epithelial cells or small clusters, indicating low-density tumour infiltration rather than solid carcinoma nests.
Reduced cellularity: Compared to adjacent tumour-rich (yellow/bright yellow) areas, the light orange regions show sparser cell populations. Tumour cells appear smaller in groups, often separated by bands of collagen.
Transitional phenotype: The morphology suggests these areas are intermediate “transition zones” between highly proliferative carcinoma and desmoplastic stroma. Some regions may represent early invasive fronts, where tumour cells begin penetrating fibrous tissue.
Possible oedematous changes: Parts of the stroma appear slightly oedematous, with looser collagen arrangement, giving a softer tissue background compared to rigid fibrosis.
So, the light orange grid areas represent stromal transition regions with loose fibrosis and scattered infiltrating carcinoma cells, reflecting a lower-density invasive front of ovarian carcinoma.

Q&A about sample tissue area for cluster-5

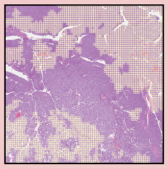

Tell me what morphological phenotype the exactly **Light Cream Yellow** grid indicated area in the tissues given to you shows?

The light cream yellow grid–marked regions in this ovarian cancer H&E section highlight a morphological phenotype with the following features:
Viable tumour epithelium with glandular/solid architecture:
These regions contain densely cellular malignant epithelial tissue, forming solid nests and irregular gland-like structures. Some tumour areas show partial glandular differentiation with compressed luminal spaces.
Cytological atypia:
The tumour cells display pleomorphic, hyperchromatic nuclei with nuclear crowding and overlapping. Prominent nucleoli and loss of polarity are evident, supporting a high-grade carcinoma phenotype.
Moderate stromal reaction: While tumour cells are dominant, there is also fibrovascular stroma present, with connective tissue interspersed between tumour nests. The stromal component is less extensive than in desmoplastic zones (e.g., bright gold or light orange areas).
Invasive growth pattern: The tumour shows irregular infiltration into surrounding tissue, with jagged borders between carcinoma and stroma. This confirms an invasive morphology rather than a confined or in-situ pattern.
In overview, the light cream yellow grid areas represent viable invasive carcinoma with solid and glandular growth, marked nuclear atypia, and a moderate stromal component, typical of high-grade serous carcinoma with active tumour proliferation.

**UNI**

Q&A about sample tissue area for cluster-5

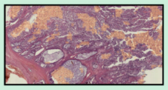

Tell me what morphological phenotype the exactly **Orange** grid indicated area in the tissues given to you shows?

The orange grid-marked regions in this ovarian cancer H&E section highlight a morphological phenotype characterised by:
Well-formed glandular/cribriform structures:
The orange-marked areas correspond to tumour epithelium forming complex gland-like or cribriform patterns. There are irregular luminal spaces, some of which are fused, giving a back-to-back gland appearance. This reflects a glandular differentiation pattern within the carcinoma.
Crowded malignant epithelium: The glands are lined by atypical epithelial cells with enlarged, pleomorphic nuclei and conspicuous nucleoli. Nuclear stratification and loss of polarity are evident, consistent with high-grade features.
Stromal infiltration: The glandular structures infiltrate into the surrounding desmoplastic stroma, confirming an invasive pattern. Some glands appear compressed by stroma, creating an irregular contour.
Focal luminal necrosis or secretions: Within certain glandular spaces, necrotic debris or eosinophilic secretions can be observed, suggesting high cell turnover.
So, the orange grid areas represent invasive carcinoma with glandular/cribriform architectural phenotype, showing malignant epithelial proliferation in irregular, back-to-back gland-like structures with nuclear atypia and stromal invasion.

Q&A about sample tissue area for cluster-6

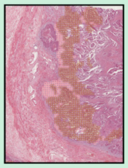

Tell me what morphological phenotype the exactly **Brown** grid indicated area in the tissues given to you shows?

The brown grid–marked regions in this ovarian cancer H&E section demonstrate a morphological phenotype with the following features:
Papillary and glandular tumour architecture: These areas show complex branching papillary structures and irregular glandular spaces lined by malignant epithelial cells. The papillae often have fibrovascular cores and project into luminal spaces.
Cytological atypia: The lining tumour cells exhibit enlarged, pleomorphic, hyperchromatic nuclei with prominent nucleoli. Nuclear stratification and crowding are evident, reflecting high-grade morphology.
Infiltrative growth pattern: The papillary and glandular elements extend into adjacent desmoplastic stroma, confirming invasive carcinoma. Some glands appear irregular and angulated, consistent with stromal invasion.
Focal luminal necrosis or secretory material: Within certain papillary and glandular spaces, eosinophilic secretions or necrotic debris may be present, indicating brisk tumour turnover.
In summary, the brown grid areas represent invasive papillary/glandular carcinoma phenotype, characterised by complex papillary projections, malignant glandular structures, marked nuclear atypia, and stromal invasion.

**Giga**

Q&A about sample tissue area for cluster-6

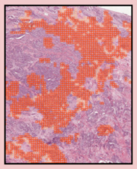

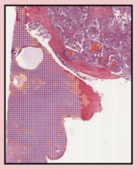

Tell me what morphological phenotype the exactly **Bright Red** grid indicated area in the tissues given to you shows?

The bright red grid–marked regions in these ovarian cancer H&E slides represent a morphological phenotype with the following characteristics:
Highly fibrotic/desmoplastic stroma: These zones are dominated by dense eosinophilic collagen bundles with elongated fibroblasts. The tissue appears homogenised and relatively acellular compared with tumour-rich areas.
Minimal epithelial tumour component: Very few malignant epithelial nests are present, often appearing as scattered or isolated cells within the fibrotic matrix. This suggests tumour regression zones or strong stromal dominance.
Reactive stromal changes: Evidence of stromal activation (desmoplasia) is present, including fibroblastic proliferation and collagen deposition. Occasional inflammatory infiltrates and vascular structures are embedded within the fibrotic background.
Transitional morphology: These bright red regions typically occur at the tumour–stroma interface, where carcinoma has either regressed or is only sparsely represented, leaving behind a fibrous tissue scaffold.
In summary:
The bright red grid areas correspond to dense desmoplastic stromal regions with minimal residual carcinoma, reflecting a stromal-dominant phenotype that may represent tumour regression or strong host stromal response.

Q&A about sample tissue area for cluster-7

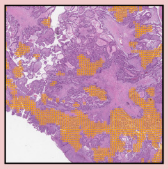

Tell me what morphological phenotype the exactly **Orange** grid indicated area in the tissues given to you shows?

The orange grid-marked regions in this ovarian cancer H&E section represent a morphological phenotype characterised by:
Papillary and glandular tumour structures: The orange-marked areas contain complex papillary fronds and irregular glandular spaces lined by malignant epithelial cells. Some papillae possess fibrovascular cores, a hallmark of papillary growth in high-grade serous carcinoma.
Cytological atypia: The epithelial lining shows pleomorphic, hyperchromatic nuclei with nuclear stratification and prominent nucleoli. Cells appear crowded and irregular, consistent with high-grade malignancy.
Invasive growth pattern: These papillary/glandular formations infiltrate into the surrounding stroma, creating jagged, irregular outlines that indicate invasion rather than in-situ growth.
Occasional luminal contents: Some glandular/papillary spaces contain eosinophilic secretions or necrotic debris, reflecting brisk tumour activity and high cell turnover.
So, the orange grid areas correspond to an invasive papillary–glandular carcinoma phenotype, characterised by complex papillary projections, glandular differentiation, marked nuclear atypia, and stromal invasion, typical of high-grade serous ovarian carcinoma.

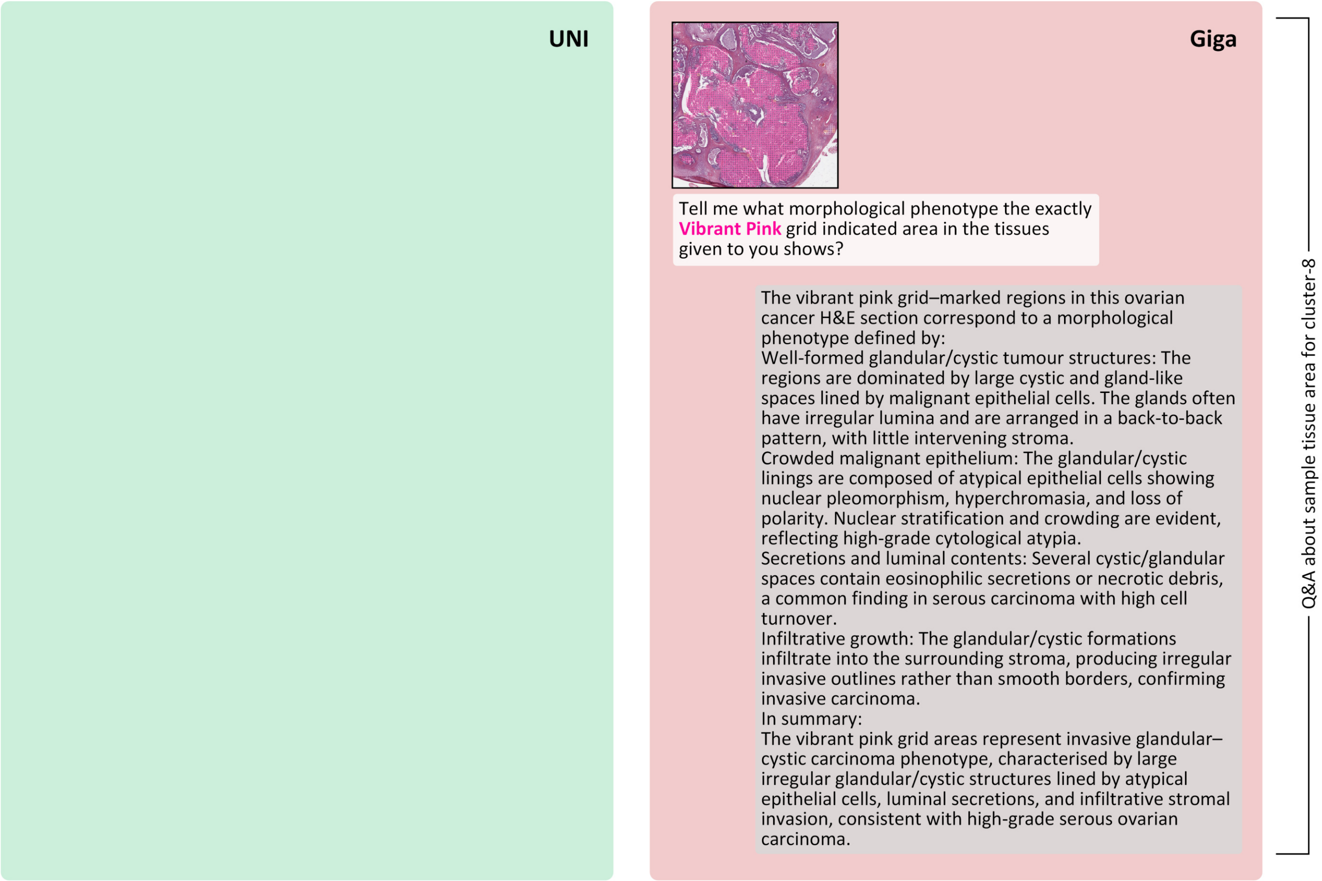


**Figure S8.** **Spatial distributions and LLM-based descriptions of attention-influx clusters (Ovarian-Bev-Resp).** Each panel contrasts UNI (*left*) and prov-GigaPath (*right*) by showing the spatial distribution of tile clusters with significant attention influx over tissue, together with morphology-focused summaries generated by a locally deployed LLM for representative clusters. Clusters were obtained via unsupervised grouping of tiles exhibiting significant attention influx, and illustrate distinct histological phenotypes prioritised by the two foundation models. The figure is divided across five pages and shares this single caption.

Initial prompt

You are a pathologist, and you need to help me review some pathology image analysis results. These images are H&E-stained pathology images from patients with ovarian cancer. I will now give you some pathological images within a local tissue range, in which boxes of different colours indicate different small areas. I will ask you about the tissue histomorphological phenotypes exhibited by 1-2 of the colour-indicated areas.

**UNI**

Q&A on reginal attention-shift examples of efflux c-1 to influx c-4

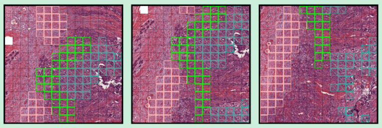

What histomorphological phenotypes are present in the regions indicated by the **Cadet Blue** and **Light Pink** boxes, respectively? What phenotypic evolution does the region undergo from **Cadet Blue** to **Light Pink**?

The Cadet Blue region shows a tumour-cell–rich phenotype, where malignant epithelial cells are densely packed in solid sheets and nests. The nuclei are enlarged and hyperchromatic, with a high nuclear-to-cytoplasmic ratio, and glandular structures are largely lost. This represents an actively proliferating and invasive area of carcinoma.
The Light Pink region is characterised by reduced tumour cellularity and a prominent stromal reaction. Dense fibrotic tissue with abundant collagen and spindle-shaped fibroblasts dominates the field, while tumour cells appear scattered and less cohesive. This reflects a fibrotic or desmoplastic phenotype with stromal overgrowth.
Together, these areas demonstrate a phenotypic evolution from a high-cellularity, tumour-dominated state (Cadet Blue) to a fibrosis-dominated, stroma-rich state (Light Pink). This shift highlights the interaction between tumour and stroma, where an invasive carcinoma front is gradually replaced or encased by reactive fibrosis.

Q&A on reginal attention-shift examples of efflux c-2 to influx c-1

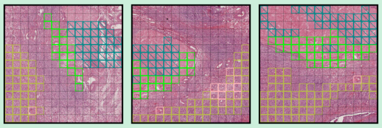

What histomorphological phenotypes are present in the regions indicated by the **Dark Cyan** and **Olive Green** boxes, respectively? What phenotypic evolution does the region undergo from **Dark Cyan** to **Olive Green**?

The Dark Cyan region shows a phenotype dominated by stromal or fibrotic tissue with relatively sparse tumour cells. The stroma appears collagenised, with pale eosinophilic extracellular matrix and scattered spindle-shaped fibroblastic elements. Tumour cells are less conspicuous, and the overall architecture reflects a fibrosis-rich or desmoplastic response zone, with cancer cells entrapped within fibrous tissue.
The Olive Green region, in contrast, demonstrates a higher density of malignant epithelial cells arranged in solid nests and sheets. The nuclei are hyperchromatic and pleomorphic, with increased nuclear-to-cytoplasmic ratio, and the tissue is overall more cellular and compact. The stromal component is present but less dominant compared with the tumour cell population, indicating a tumour-rich phenotype.
The evolution from Dark Cyan to Olive Green represents a transition from a fibrosis-dominated, stroma-rich microenvironment to a cellularity-rich, tumour-dominated phenotype. This shift highlights how areas of desmoplastic stroma can give way to expanses of actively proliferating carcinoma, reflecting dynamic tumour–stroma interactions and the spatial heterogeneity of the tumour bed.

**Giga**

Q&A on reginal attention-shift examples of efflux c-2 to influx c-3

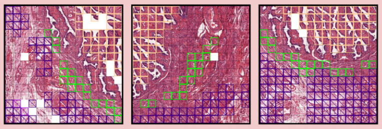

What histomorphological phenotypes are present in the regions indicated by the **Indigo** and **Light Orange** boxes, respectively? What phenotypic evolution does the region undergo from **Indigo** to **Light Orange**?

The Indigo region is characterised by dense fibrous stroma with abundant collagen deposition and elongated spindle-shaped stromal cells. The architecture is dominated by fibrotic tissue with relatively few viable tumour epithelial elements. Blood vessels and stromal remodelling are also apparent, giving the area a desmoplastic and stromal-rich phenotype.
The Light Orange region, in contrast, is composed of well-formed malignant epithelial structures. Tumour cells form glandular and papillary architectures with prominent nuclear atypia and stratification. The cellular density is higher, and epithelial proliferation is the dominant feature, with less intervening stroma compared to the Indigo zone.
The evolution from Indigo to Light Orange represents a transition from a stromal-rich, fibrotic/desmoplastic phenotype to a tumour-cell–dominant, glandular/papillary epithelial phenotype. This reflects the spatial heterogeneity of ovarian carcinoma, where areas of stromal reaction and fibrosis (Indigo) give way to zones of active epithelial tumour growth (Light Orange), illustrating the dynamic interplay between tumour cells and their microenvironment.

Q&A on reginal attention-shift examples of efflux c-7 to influx c-0

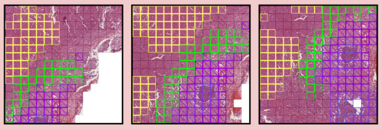

What histomorphological phenotypes are present in the regions indicated by the **Dark Violet** and **Bright Yellow** boxes, respectively? What phenotypic evolution does the region undergo from **Dark Violet** to **Bright Yellow**?

The Dark Violet region is dominated by stromal and necrotic/haemorrhagic components. It shows extensive eosinophilic stroma, vascular congestion, and areas of necrosis with reduced viable tumour cell density. The architecture is less defined, reflecting stromal remodelling, tissue degeneration, and haemorrhagic changes rather than organised tumour growth.
The Bright Yellow region, in contrast, displays a tumour-rich phenotype with dense sheets and nests of malignant epithelial cells. The nuclei are enlarged, hyperchromatic, and pleomorphic, with high nuclear-to-cytoplasmic ratios. The stroma is minimal, and the carcinoma cells are the dominant component, reflecting an actively proliferating tumour phenotype.
The phenotypic evolution from Dark Violet to Bright Yellow represents a shift from a stromal- and necrosis-dominated environment towards a cellularity-rich, viable carcinoma phenotype. This highlights tumour heterogeneity, where regions of stromal reaction and degeneration transition into areas of aggressive, viable epithelial tumour growth.

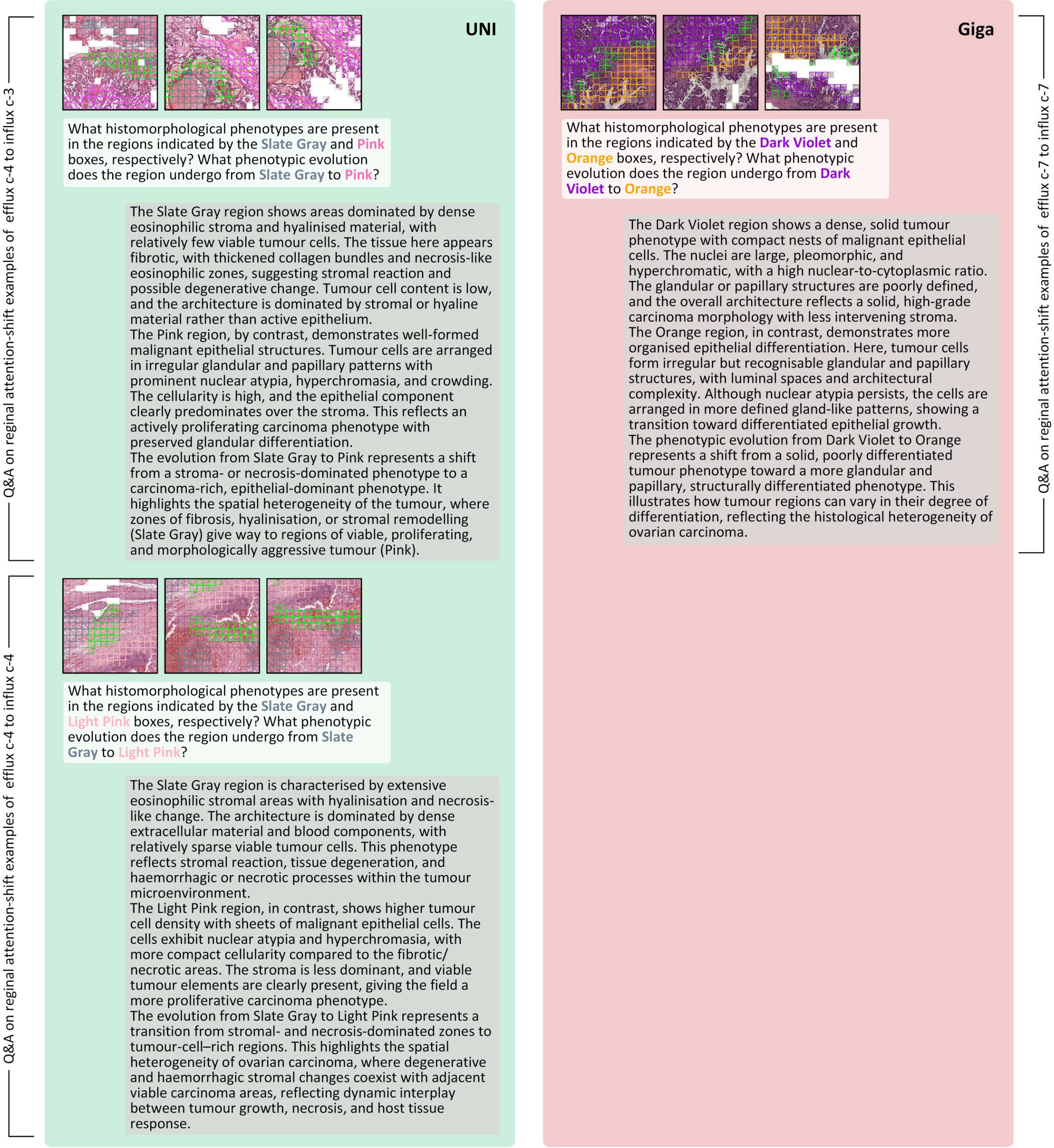


**Figure S9. LLM-based descriptions of regional attention-shift patterns (Ovarian-Bev-Resp).** Examples of local attention-shift patterns based on UNI and prov-GigaPath embeddings in the Ovarian-Bev-Resp dataset. After the initial prompt, each panel shows the locally deployed LLM's generative dialogue records on histomorphological phenotypes across efflux-to-influx transitions. The left panels correspond to UNI, and the right panels to prov-GigaPath. The figure is divided across two pages and shares this single caption.

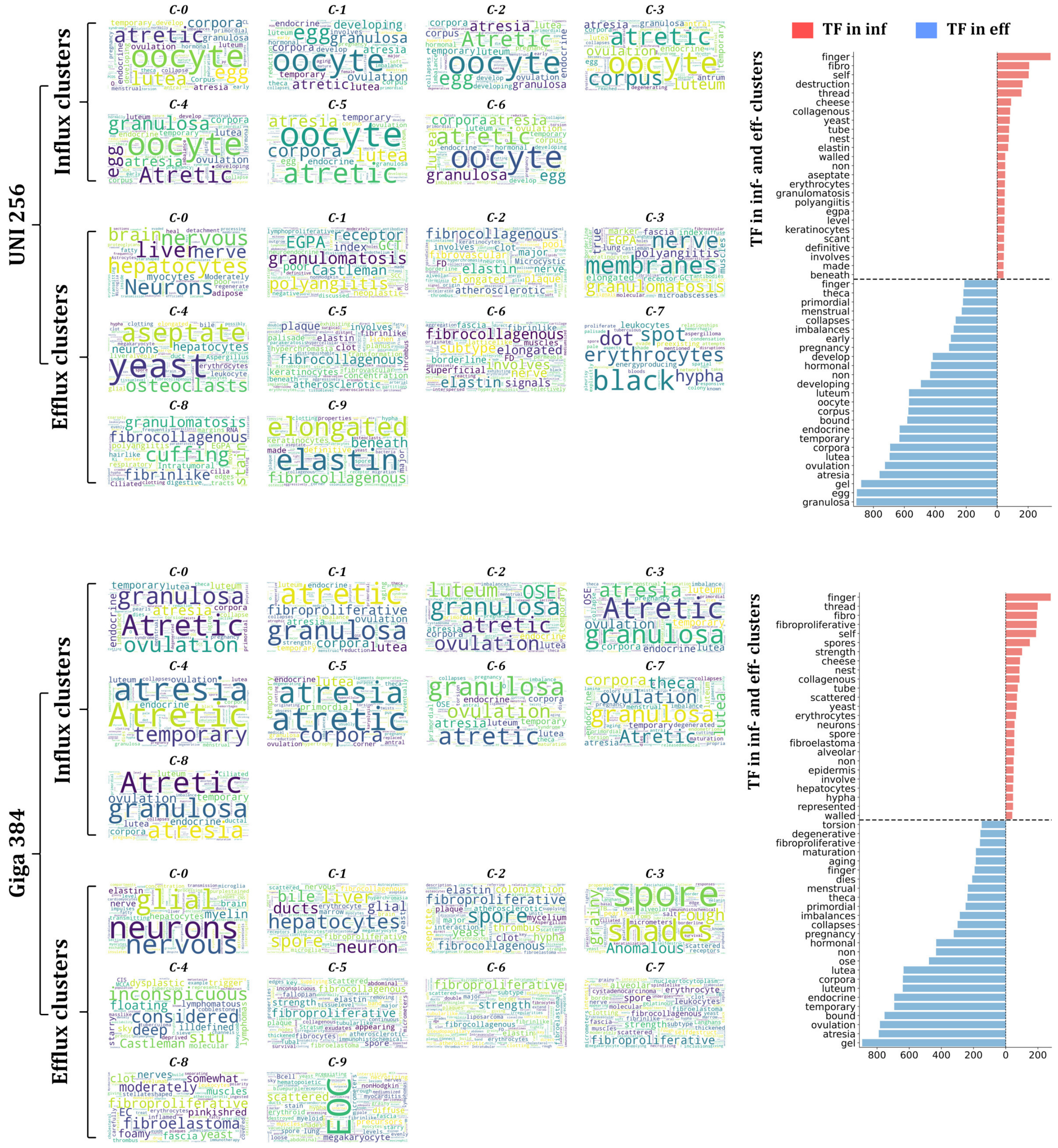


**Figure S10. Word cloud and term frequency analysis of local LLM-generated descriptions in Ovarian-Bev-Resp.** For each cluster of tiles with significant attention influx or efflux, a medical-specific small-scale LLM was used to generate concise histopathological descriptions. The resulting terms were aggregated to construct word clouds (left) and term frequency (TF) plots (right). Results are shown separately for UNI and prov-GigaPath.

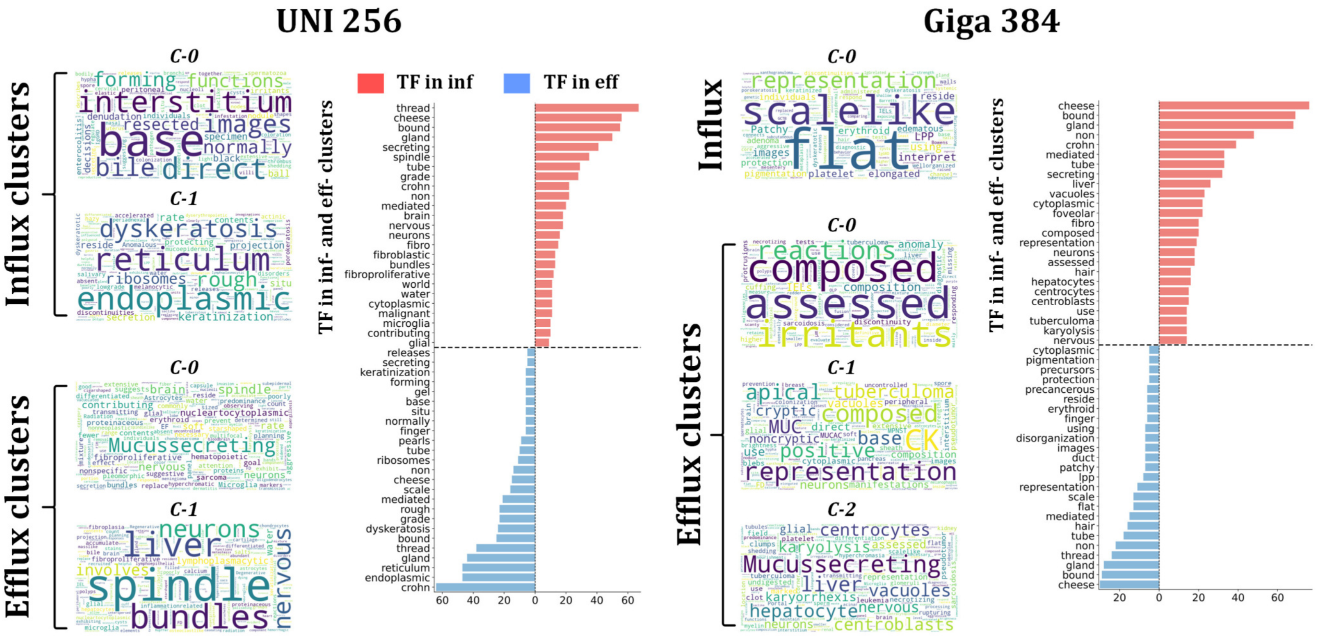


**Figure S11.** **Word cloud and term frequency analysis of local LLM-generated descriptions in FOCUS-KRAS.** Clusters of tiles exhibiting attention influx or efflux were summarised using a medical-specific LLM, and their outputs visualised as word clouds and term frequency plots. Results are shown separately for UNI and prov-GigaPath.

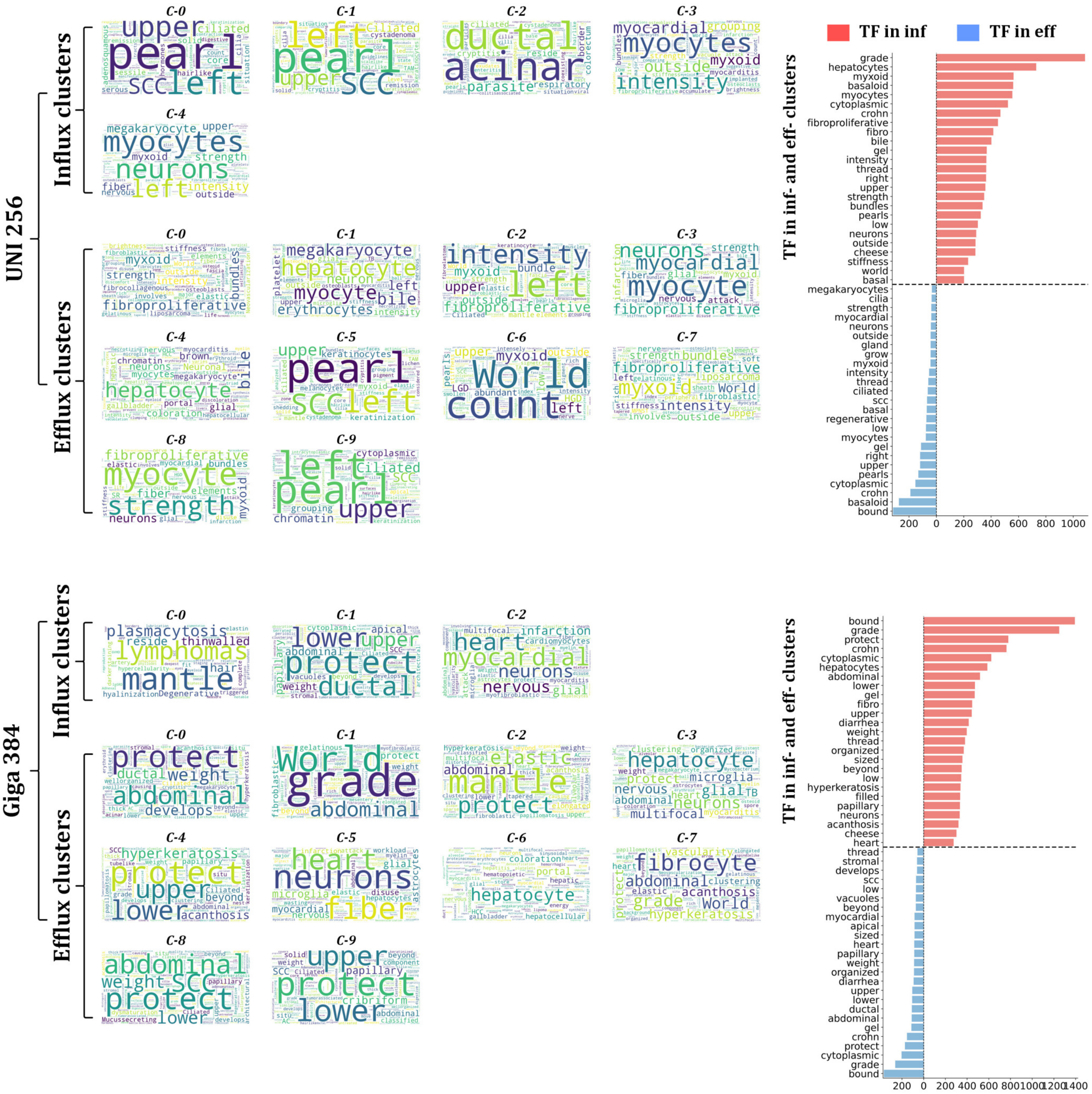


**Figure S12. Word cloud and term frequency analysis of local LLM-generated descriptions in Colitis-Marsh.** Representative influx and efflux clusters were annotated using a medical-specific LLM, with outputs visualised as word clouds and term frequency plots. Results are shown separately for UNI and prov-GigaPath.
